# Anti-PEG Antibodies Boosted in Humans by SARS-CoV-2 Lipid Nanoparticle mRNA Vaccine

**DOI:** 10.1101/2022.01.08.22268953

**Authors:** Yi Ju, Wen Shi Lee, Emily H. Pilkington, Hannah G. Kelly, Shiyao Li, Kevin J. Selva, Kathleen M. Wragg, Kanta Subbarao, Thi H.O. Nguyen, Louise C. Rowntree, Lilith F. Allen, Katherine Bond, Deborah A. Williamson, Nghia P. Truong, Magdalena Plebanski, Katherine Kedzierska, Siddhartha Mahanty, Amy W. Chung, Frank Caruso, Adam K. Wheatley, Jennifer A. Juno, Stephen J. Kent

**Affiliations:** Department of Microbiology and Immunology, Peter Doherty Institute for Infection and Immunity, The University of Melbourne, Melbourne, VIC 3000, Australia; School of Health and Biomedical Sciences, RMIT University, Bundoora, VIC 3083, Australia; Department of Chemical Engineering, The University of Melbourne, Melbourne, VIC 3000, Australia; Department of Drug Delivery, Disposition and Dynamics, Monash Institute of Pharmaceutical Sciences, Monash University, Melbourne, VIC 3000, Australia; WHO Collaborating Centre for Reference and Research on Influenza, Peter Doherty Institute for Infection and Immunity, Melbourne, VIC 3000, Australia; Department of Microbiology, Royal Melbourne Hospital, Melbourne, VIC 3000, Australia; Department of Infectious Diseases, Peter Doherty Institute for Infection and Immunity, The University of Melbourne, Melbourne, VIC 3000, Australia; Melbourne Sexual Health Centre and Department of Infectious Diseases, Alfred Hospital and Central Clinical School, Monash University, Melbourne, VIC 3000, Australia

**Keywords:** PEGylated lipid nanoparticle, COVID-19, immunoglobulins, biomolecular coronas, human blood assay, particle–immune cell interactions

## Abstract

Humans commonly have low level antibodies to poly(ethylene) glycol (PEG) due to environmental exposure. Lipid nanoparticle (LNP) mRNA vaccines for SARS-CoV-2 contain small amounts of PEG but it is not known whether PEG antibodies are enhanced by vaccination and what their impact is on particle–immune cell interactions in human blood. We studied plasma from 130 adults receiving either the BNT162b2 (Pfizer-BioNTech) or mRNA-1273 (Moderna) mRNA vaccines, or no SARS-CoV-2 vaccine for PEG-specific antibodies. Anti-PEG IgG was commonly detected prior to vaccination and was significantly boosted a mean of 13.1-fold (range 1.0 to 70.9) following mRNA-1273 vaccination and a mean of 1.78-fold (range 0.68 to 16.6) following BNT162b2 vaccination. Anti-PEG IgM increased 68.5-fold (range 0.9 to 377.1) and 2.64-fold (0.76 to 12.84) following mRNA-1273 and BNT162b2 vaccination, respectively. The rise in PEG-specific antibodies following mRNA-1273 vaccination was associated with a significant increase in the association of clinically relevant PEGylated LNPs with blood phagocytes *ex vivo*. PEG antibodies did not impact the SARS-CoV-2 specific neutralizing antibody response to vaccination. However, the elevated levels of vaccine-induced anti-PEG antibodies correlated with increased systemic reactogenicity following two doses of vaccination. We conclude that PEG-specific antibodies can be boosted by LNP mRNA-vaccination and that the rise in PEG-specific antibodies is associated with systemic reactogenicity and an increase of PEG particle–leukocyte association in human blood. The longer-term clinical impact of the increase in PEG-specific antibodies induced by lipid nanoparticle mRNA-vaccines should be monitored.

---

Humans are exposed to poly(ethylene) glycol (PEG) through consumer products and medicines. A proportion of adults have circulating anti-PEG antibodies.^1-5^ Incorporation of PEG into the formulation of some systemically administered drugs to improve pharmacokinetics can result in anti-PEG antibodies that ultimately limit the bioavailability of the drug and can cause side effects.^6-8^ Lipid nanoparticle (LNP) mRNA vaccines for SARS-CoV-2 contain small amounts of a PEG-lipid conjugate to stabilize the LNPs.^9,10^ An estimated 50 µg and 117 µg (calculated based on total lipid dose and molar lipid ratios^11^) of the PEG-lipid conjugates ALC-0159 and PEG2000-DMG are present per dose for the BNT162b2 (Comirnaty, Pfizer-BioNTech) and mRNA-1273 (Spikevax, Moderna) vaccines, respectively.^12,13^ Over a billion doses of SARS-CoV-2 LNP mRNA vaccines containing PEG-lipids have now been given worldwide. However, it is not known whether anti-PEG antibodies are induced or boosted by intramuscular LNP mRNA vaccination for COVID-19 and what effect, if any, this may have on vaccine responses.

We studied serial blood samples from 130 adults across 3 separate cohorts, including 55 adults receiving 2 doses of BNT162b2 vaccines (subject details in Table S1), 20 adults receiving 2 doses of mRNA-1273 vaccines (subject details in Table S2), and 55 unvaccinated subjects (including 40 SARS-CoV-2 infected convalescent patients and 15 healthy donors, subject details in Table S3). PEG-specific IgG and IgM antibodies were quantified by ELISA first to a 40 kilodalton PEG molecule as described in the Methods. For BNT162b2 and mRNA-1273 cohorts, PEG-specific IgG was detectable (endpoint titre >1:10) prior to vaccination in 53 of the 75 subjects (71%), ranging in titre from 1:12-1:3000. Of the 75 subjects, 31 subjects (41%) had a titre >1:100. This is consistent with the wide range of prevalence of anti-PEG antibodies previously reported, ranging from <4% to 72%,^3-5^ potentially attributed to differences in geographic location, age of the subjects as well as different assay systems used in the individual studies. A small but consistent increase in PEG-specific IgG and IgM was observed after 2 doses of BNT162b2 vaccinations with a mean fold change of 1.78 (range 0.68−16.6) and 2.64 (range 0.76−12.84), respectively (Figure 1A). There were 9 (16%) or 23 (42%) subjects who had a boost of anti-PEG IgG or IgM with more than 2-fold change after BNT162b2 vaccination, respectively. Only 1 or 2 subjects had a boost of anti-PEG IgG or IgM with more than 10-fold change after BNT162b2 vaccination, respectively. A time course analysis showed that subjects with increased PEG antibody responses had a stepwise increase after the first and second doses of BNT162b2 vaccination (Figure S4). PEG antibodies did not increase further out to 3 months in a subset of subjects (*n* = 10) where late post-vaccination samples were available (Figure S4B). A much larger increase in anti-PEG IgG and IgM was observed after 2 doses of mRNA-1273 vaccinations with a mean fold change of 13.1 (range 1.0−70.9) and 68.5 (range 0.9−377.1), respectively (Figure 1B). There were 15 (75%) or 18 (90%) subjects who had a boost of anti-PEG IgG or IgM with more than 2-fold change after mRNA-1273 vaccination, respectively. Six (30%) or 10 (50%) subjects had a boost of anti-PEG IgG or IgM with more than 10-fold change after mRNA-1273 vaccination, respectively. In a control group of 55 unvaccinated subjects, anti-PEG IgG and IgM titres did not increase and overall slightly decreased over a 6-month period with a mean fold change of 0.92 and 0.98, respectively (Figure 1C). Comparing the fold change of anti-PEG antibody levels across the 3 cohorts (Figure 1D), we concluded that both anti-PEG IgG and IgM were significantly boosted by mRNA-1273 vaccination and modestly boosted by BNT162b2 vaccination. The difference in PEG immunogenicity between two vaccines might be due to a higher dose of PEGylated lipid in the mRNA-1273 vaccine (117 µg vs 50 µg PEG-lipid per dose) compared to the BNT162b2 vaccine. PEG immunogenicity is affected by the terminal group of PEG in that methoxy-PEG is more immunogenic than hydroxy-PEG^14^, although the PEG-lipids in the mRNA-1273 (PEG2000-DMG) and BNT162b2 formulation (ALC-0159) both contain a methoxy-PEG2000. Furthermore, the boost of anti-PEG antibody in mice depends on the rate of PEG-lipid shedding off the LNP.^15^ Future animal studies are required to determine the impact of PEG-lipid structure and LNP formulation on PEG immunogenicity of mRNA LNP vaccines.

**Figure 1.**
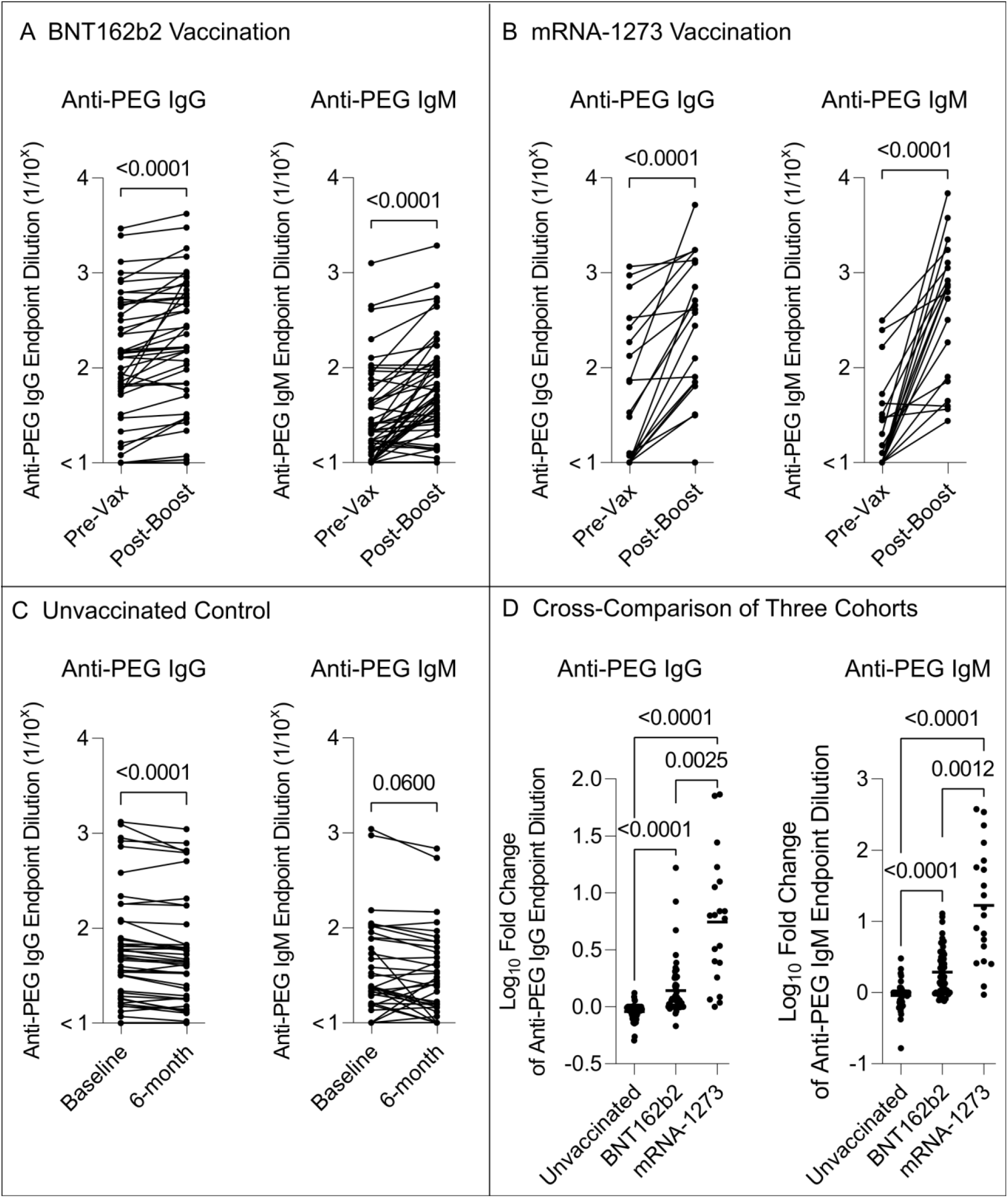
The impact of SARS-COV-2 LNP mRNA vaccination on the levels of PEG specific antibodies. A) Comparing plasma anti-PEG IgG and IgM titres before vaccination (Pre-Vax) and 2 to 7 weeks (mean 27 days, range 12−49) after second dose (Post-Boost) of the BNT162b2 vaccination of healthy cohort (n=55, see raw data in Figure S1). B) Comparing plasma anti-PEG IgG and IgM titres before vaccination (Pre-Vax) and 3 weeks (mean 19 days, range 18−27) after second dose (Post-Boost) of the mRNA-1273 vaccination of healthy cohort (n=20, see raw data in Figure S2). C) Comparing plasma anti-PEG IgG and IgM titres over a 6-month period (mean 186 days, range 120−253) of unvaccinated control cohort (n=55, including 40 SARS-CoV-2 infected convalescent patients and 15 healthy donors, see raw data in Figure S3). D) Cross-comparison of the fold change (log_10_) of anti-PEG IgG and IgM titres among three cohort: BNT162b2 (Post-Boost/Pre-Vax, n=55), mRNA-1273 (Post-Boost/Pre-Vax, n=20), and unvaccinated control cohort (6-month/baseline, n=55). p-values in (A−C) were derived by Wilcoxon’s matched-pairs signed rank test and in (D) were derived by nonparametric Kruskal-Wallis test with Dunn’s multiple comparisons test.

We observed a sex and age-dependent PEG-specific antibody levels in 130 subjects across 3 cohorts. Females had higher titres of pre-existing anti-PEG IgG (mean titre 1:73 vs. 1:35) and IgM (mean titre 1:28 vs. 1:21) than males (Figure 2A). This is consistent with previous findings and has been postulated to be related to greater exposure of females to PEG-containing cosmetic products.^3^ We found the age of subjects was negatively correlated with the pre-existing levels of anti-PEG IgG and IgM (Figure 2B). Younger subjects have been previously reported to have higher PEG-specific antibodies in a large serosurvey.^3^ However, we did not observe an influence of gender or age on the change in anti-PEG IgG or IgM after BNT162b2 or mRNA-1273 vaccination of 75 subjects (Figure 2C,D).

**Figure 2.**
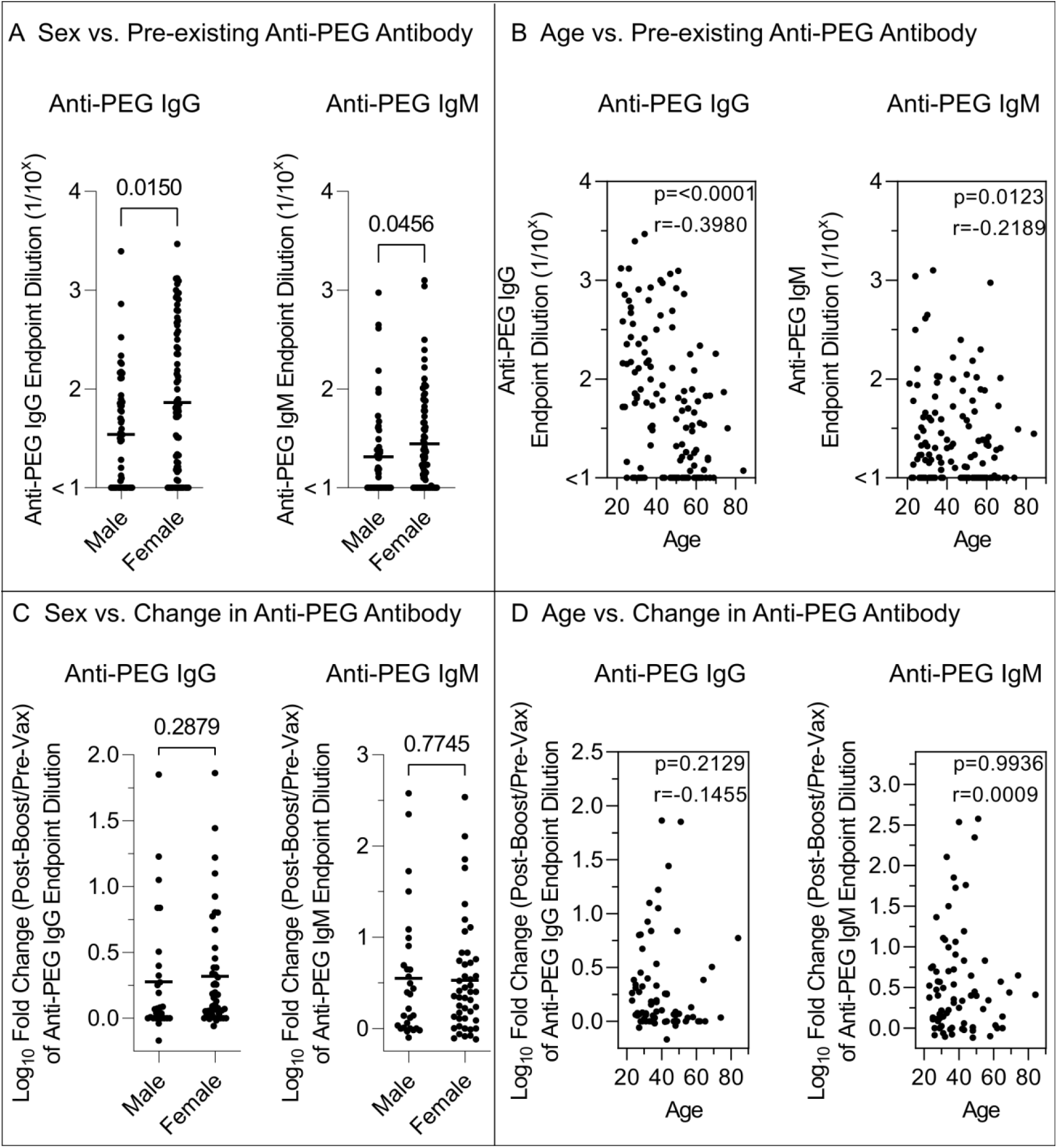
The impact of Sex and Age on the levels of PEG-specific antibodies. A) Comparing the pre-existing (Pre-Vax) anti-PEG IgG or IgM titres between male and female donors (including all the vaccinated and unvaccinated control cohorts, *n* = 130, 53 male vs. 77 female). B) Spearman correlation between age of donors and the pre-existing (Pre-Vax) anti-PEG IgG or IgM titres (including all the vaccinated and unvaccinated control cohorts, *n* = 130). C) Comparing the fold change (log_10_) of anti-PEG IgG or IgM titres (Post-Boost/Pre-Vax) between male and female donors after 2 doses of vaccination (including all the BNT162b2 and mRNA-1273 cohorts, n=75, 29 male vs. 46 female). D) Spearman correlation between age of donors and the fold change (log_10_) of anti-PEG IgG or IgM titres (Post-Boost/Pre-Vax) after 2 vaccinations, including all the BNT162b2 and mRNA-1273 cohorts (*n* = 75). p-values in (A,C) were derived by Mann-Whitney *U* test and in (B,D) were derived by Spearman correlation analysis.

To determine whether the PEG antibodies detected specifically recognized the PEG-lipid or other synthetic lipid components of the BNT162b2 vaccine, we assessed a subset of 18 vaccinees where larger plasma samples were available for antibody response to the ALC-0159 (PEG-lipid), ALC-0315 (ionizable lipid), and DSPC (helper lipid) components of the BNT162b2 vaccine. We found anti-human IgG that was specific to the ALC-0159 PEG-lipid increased in 10 out of 18 subjects studied (mean fold change 3.03, range 1−27.4, Figure 3A). The increase of PEG-lipid-specific IgG correlated with the increase of PEG-specific IgG from the same subjects detected using a 40 kilodalton PEG (Figure 3B). In contrast, the other lipid components, ALC-0315 and DSPC, elicited no antibody responses before or after vaccination (Figure 3C). Furthermore, a humanized monoclonal anti-PEG IgG was used to assess specificity to the synthetic lipid components of BNT162b2 vaccine. We found the monoclonal anti-PEG IgG specifically recognized the ALC-0159 (PEG-lipid) and 40 kilodalton PEG in a concentration-dependent manner, whilst it did not recognize ALC-0315 or DSPC (Figure 3D). Taken together, these results suggest antibody recognition of PEG in the PEG-lipid component of the BNT162b2 vaccine occurs in vaccinees.

**Figure 3.**
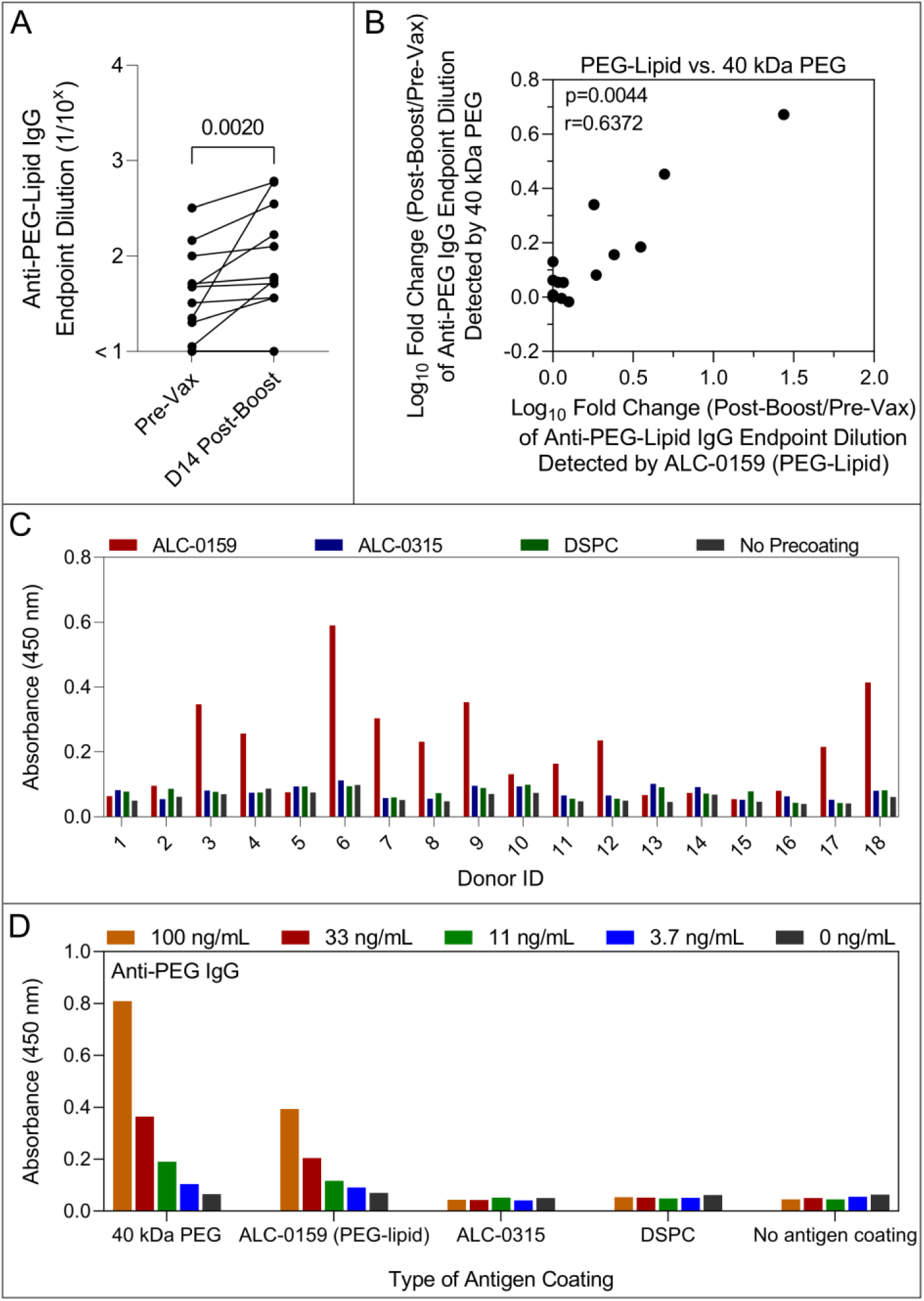
Specificity of PEG specific antibodies to PEG-lipid and other synthetic lipid components of BNT162b2 vaccine. A) PEG-lipid-specific IgG titres before and day 14 following 2 doses of the BNT162b2 vaccination (*n* = 18, donor 1−18 from BNT162b2 vaccinee cohort). The ELISA were performed by coating ALC-0159 (PEG-lipid contained in the BNT162b2 vaccine) on hydrophobic plates. p-values were derived by Wilcoxon’s matched-pairs signed rank test. B) Spearman correlation between the fold change (log_10_) of anti-PEG-lipid IgG titres detected by ALC-0159 and the fold change of anti-PEG IgG titres detected by 40 kilodalton PEG at day 14 post-boost BNT162b2 vaccination compared to pre-vax (n=18, donor 1−18 from BNT162b2 vaccinee cohort). C) Specificity of anti-human IgG to ALC-0159 (PEG-lipid), ALC-0315 (ionizable lipid), and DSPC (helper lipid) in comparison to background (no lipid precoating) of plasma from 18 healthy subjects (donor 1−18) at day 14 post-boost BNT162b2 vaccination. The ELISA were performed by coating individual lipid components (ALC-0159, ALC-0315, and DSPC) on hydrophobic plates and the plasma were diluted at 1:10 in 5% skim milk. Data are shown as mean of two independent measurements. D) Specificity of monoclonal anti-PEG IgG to 40 kilodalton (kDa) PEG, ALC-0159, ALC-0315, and DSPC, which were pre-coated as antigen on hydrophobic ELISA plates. Data are shown as mean of two independent measurements.

PEG is a hydrophilic compound commonly formulated with therapeutics to reduce non-specific immune clearance. Anti-PEG antibodies can reduce the bioavailability and efficacy of PEG-containing therapeutics.^6^ To determine the potential of anti-PEG antibodies induced by SARS-CoV-2 LNP mRNA vaccination to promote immune-mediated clearance of nanomaterials containing PEG, we used a previously developed *ex vivo* human blood assay to assess the impact of plasma on particle−immune cell association (Experimental design shown in Scheme 1).^16^ The assay incubates nanoparticles for 1 hr with fresh primary human blood cells containing monocytes, granulocytes, and B cells in the presence of plasma before and after vaccination and measures nanoparticle and immune cell association by flow cytometry (details in Methods, gating strategy shown in Figure S5). We studied three PEGylated nanoparticles, including 100 nm PEGylated mesoporous silica (PEG-MS) nanoparticles, PEGylated doxorubicin-encapsulated liposomes that contain the same lipid composition and drug/lipid ratio as clinically used liposomal doxorubicin agent (Doxil) for anti-cancer chemotherapy,^17^ and PEGylated LNPs that were formulated with the same molar composition of lipids used in the US Food and Drug Administration-approved Onpattro (Patisiran) LNP formulation for treating transthyretin-mediated amyloidosis (characterization in Figure 4, Table S4).^18^ Both Doxil and Onpattro LNPs are intravenously administered and therefore relevant nanomedicine models for studying bio−nano interactions in human blood. All three nanoparticles displayed a range of plasma donor-dependent association with immune cells (Figures S6−8), consistent with our previous findings.^19^ We observed that the association of PEGylated nanoparticles with human blood immune cells strongly correlated with the baseline levels of anti-PEG IgG and IgM. Specifically, the association of Onpattro LNPs with granulocytes and monocytes correlated with pre-existing anti-PEG IgG and IgM (Figure 5A,B). Similarly, the association of Doxil and PEG-MS nanoparticles with monocytes and granulocytes both correlated with the pre-existing levels of anti-PEG IgG, while the association of Doxil and PEG-MS nanoparticles with B cells correlated with pre-existing anti-PEG IgM and IgG, respectively (Figures S9, S11).

**Figure 4.**
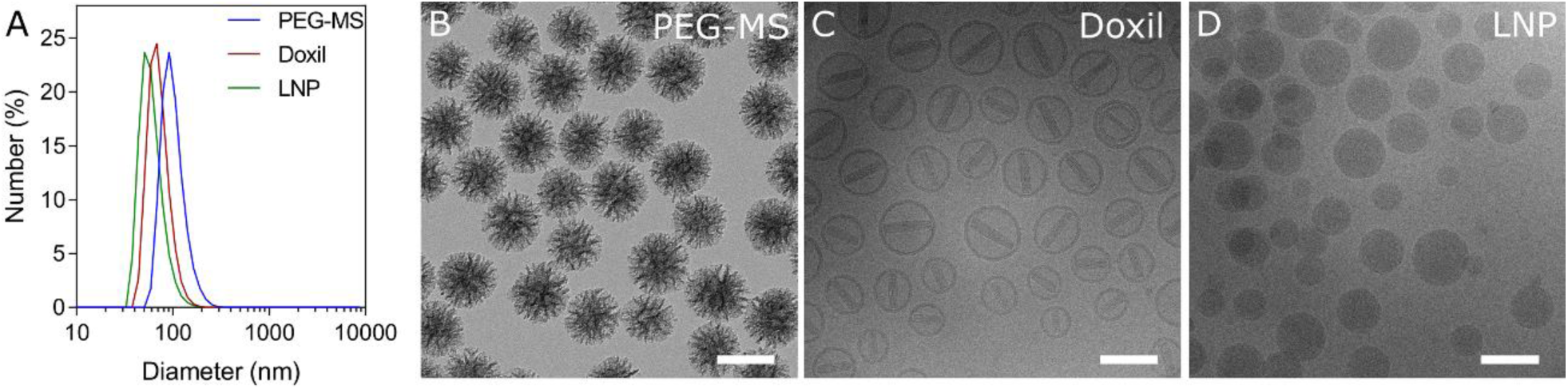
Characterization of PEGylated nanoparticles. A) Dynamic light scattering of PEG-MS nanoparticles, Doxil, and Onpattro LNPs. B) Transmission electron microscopy images of PEG-MS nanoparticles. C,D) Cryogenic transmission electron microscopy images of Doxil and LNPs. Scale bars are 100 nm in (B-D).

**Figure 5.**
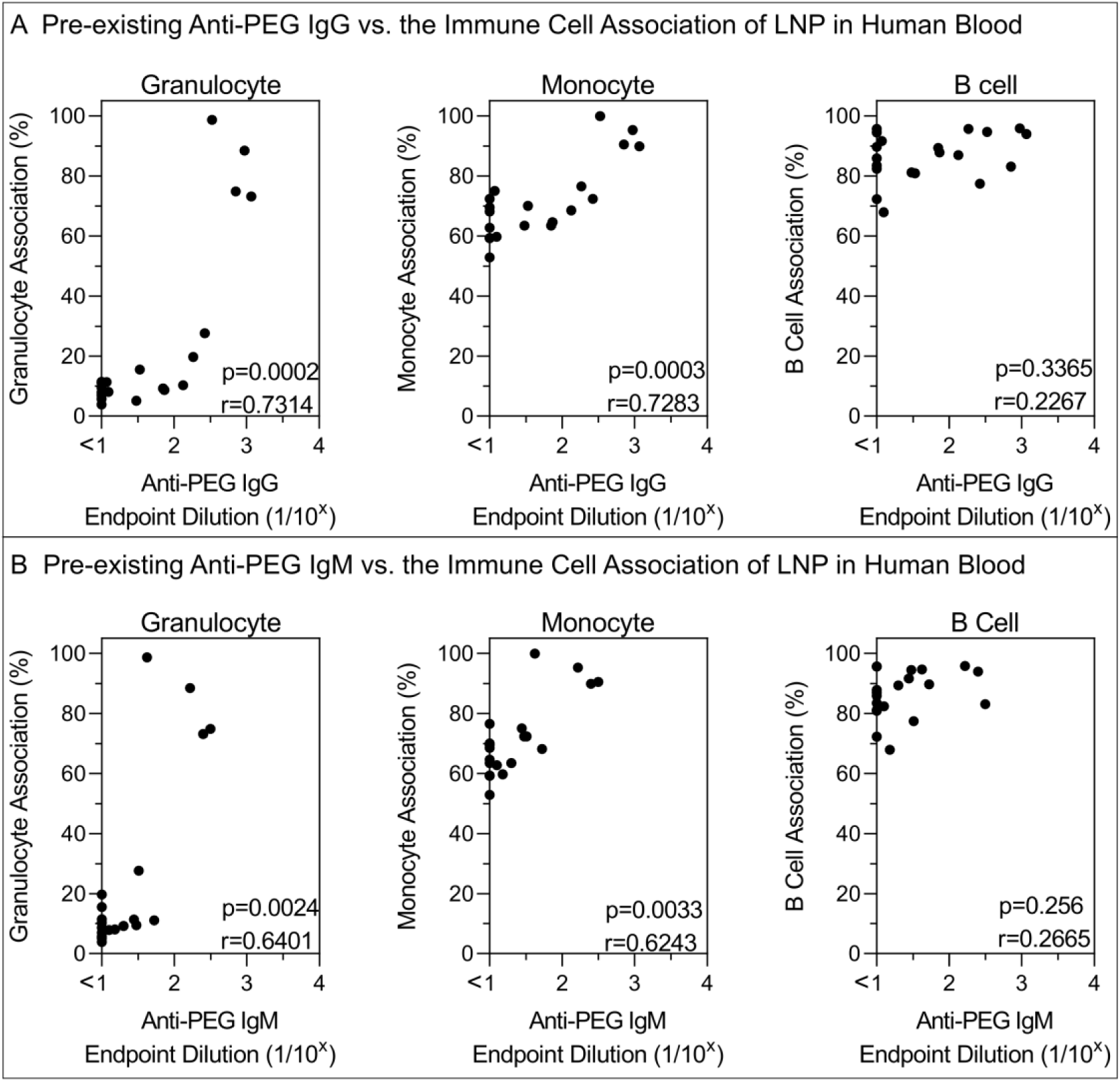
Impact of pre-existing anti-PEG antibody levels on the association of Onpattro LNPs with immune cells in human blood. A,B) Spearman correlation between pre-existing (Pre-Vax) anti-PEG IgG or IgM titres (n=20, donor 56−75 from mRNA-1273 vaccinee cohort) and LNP association with monocytes, granulocytes, or B cells in human blood. Cell association (%) refers to the proportion of each cell type with positive fluorescence, above background, stemming from fluorescence-labeled particles (see gating strategy in Figure S5.). Cell association (%) data are shown as the mean of three independent experiments (using the same batch of plasma from each donor), with at least 150,000 leukocytes analyzed for each experimental condition studied (see raw data in Figure S6).

Importantly, we observed a significant increase in the association of Onpattro LNPs with blood phagocytes (*i*.*e*., granulocytes and monocytes) in plasma obtained after mRNA-1273 vaccination (Figure 6A). The increase in anti-PEG IgG and IgM antibodies after mRNA-1273 vaccination correlated with the increase in association of Onpattro LNPs to granulocytes and monocytes (Figure 6B). Similar to Onpattro LNPs, we observed an increase in Doxil−monocyte association and PEG-MS nanoparticle−B cell association mediated by plasma after BNT162b2 vaccination (Figures S10A, S12A), which correlated with the level of changes in anti-PEG IgM levels in plasma (Figures S10B, S12B). The nanoparticle−B cell association is likely mediated by complement receptors on B cells since heat inactivation of plasma to destroy complement activity has previously been shown to inhibit this association.^20^ In contrast to mRNA-1273 vaccination, the increase in blood immune cell association with PEGylated nanoparticles was lower for most plasma after BNT162b2 vaccination (Figures S10, S12), likely due to the smaller boost of anti-PEG antibodies following BNT162b2 vaccination. These *ex vivo* results across three independent particle systems and two mRNA LNP vaccinee cohorts demonstrate anti-PEG antibodies boosted by mRNA LNP vaccination can increase immune cell interactions with other PEG-based nanomaterials. Further, the increased level of anti-PEG antibodies induced by mRNA-1273 vaccination significantly enhanced human blood immune cell association of the PEG-containing therapeutic Onpattro. Plasma proteins are known to opsonize foreign nanomaterials in the blood, forming so-called “biomolecular coronas”,^21-23^ which leads to recognition and inactivation of nanomaterials by the immune system.^24-27^ The formation of biomolecular coronas on Doxil *in vivo* in human systemic circulation has been previously described.^22^ Recently, the composition of biomolecular coronas was also found to regulate fate and/or utility of LNPs.^28,29^ In our previous studies we found that the enrichment of immunoglobulins and complement proteins in biomolecular coronas is correlated with donor-specific nanoparticle association with human blood immune cells.^19^ The studies herein demonstrate that anti-PEG antibodies are key immunoglobulins that drive the donor-dependent immune cell association of PEG-containing nanoparticles in human blood.

**Figure 6.**
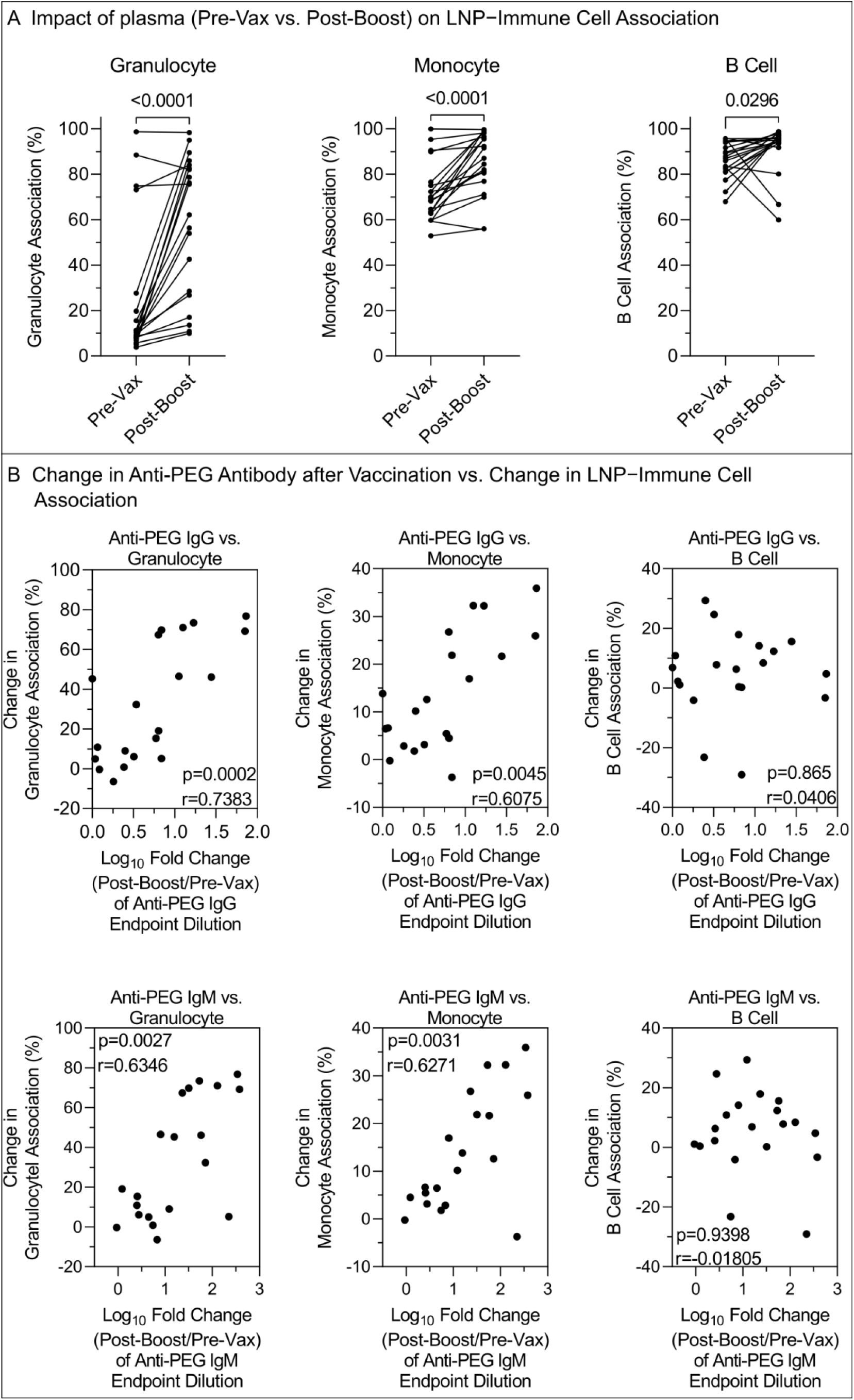
Impact of change in anti-PEG antibody after mRNA-1273 vaccination on the association of Onpattro LNPs with immune cells in human blood. A) Comparing the impact of plasma collected before vaccination (Pre-Vax) and post-boost of the mRNA-1273 vaccination (n=20, donor 56−75) on Onpattro LNP association with monocytes, granulocytes, or B cells in human blood. p-values were derived by Wilcoxon’s matched-pairs signed rank test. B) Spearman correlation between the fold change (log_10_) of anti-PEG IgG or IgM titres (Post-Boost/Pre-Vax) after 2 doses of mRNA-1273 vaccination (n=20) and change in Onpattro LNP association with monocytes, granulocytes, or B cells in human blood. Cell association (%) refers to the proportion of each cell type with positive fluorescence, above background, stemming from fluorescence-labeled particles. Cell association (%) data are shown as the mean of three independent experiments (using the same batch of plasma from each donor), with at least 150,000 leukocytes analyzed for each experimental condition studied (see raw data in Figure S6). Change in cell association (%) refers to post-boost cell association (%) minus pre-vax cell association (%).

Studies have shown the enrichment of immunoglobulins in biomolecular coronas can trigger complement activation and opsonization of nanoparticles.^30^ To understand the role of anti-PEG antibody in complement deposition, we developed an antibody-dependent complement deposition assay to assess anti-PEG antibody-mediated complement component 1q (C1q) binding in plasma collected before and after mRNA-1273 vaccination (see method section). We found that anti-PEG antibody-mediated C1q binding was negligible in plasma collected before vaccination but increased to various levels in plasma collected after 2 doses of mRNA-1273 vaccination from 11 out of 20 subjects (Figure 7A). The level of C1q binding significantly correlated with levels of anti-PEG IgG and IgM (Figure 7B), illustrating anti-PEG antibody-dependent C1q binding.

**Figure 7.**
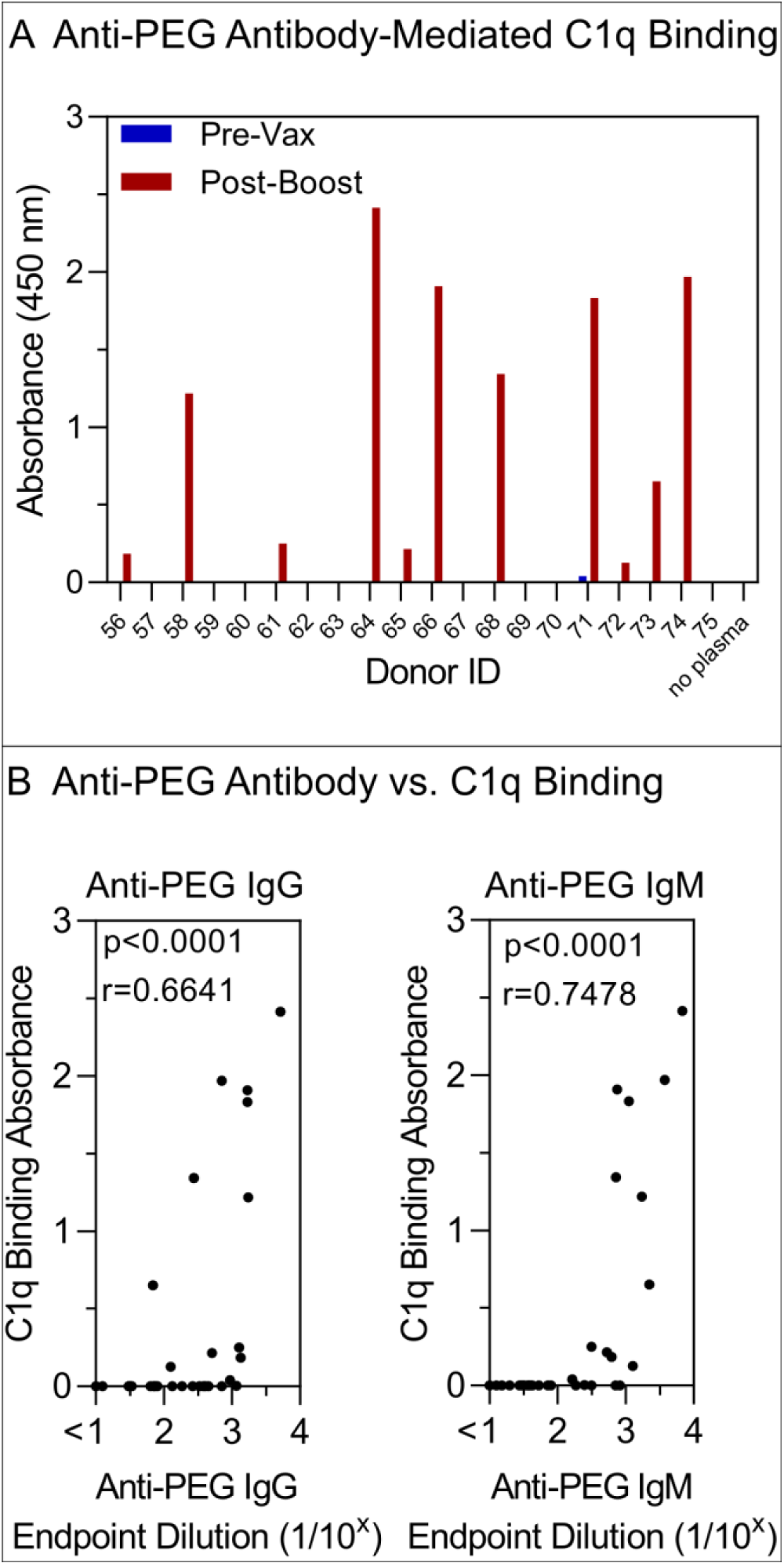
Anti-PEG antibody-mediated C1q binding. A) ELISA absorbance showing the PEG-specific antibody-mediated C1q binding of plasma collected before vaccination (Pre-Vax) and post-boost of the mRNA-1273 vaccination (n=20, donor 56−75). The ELISA were performed by coating 40kDa PEG on ELISA plates and the plasma were diluted at 1:5 in 1% bovine serum albumin (BSA), followed by washing and incubating with biotinylated C1q and horseradish peroxidase (HRP)-conjugated streptavidin. Non-PEG-precoating wells were treated the same as PEG-precoated wells and used as background for absorbance subtraction (see raw absorbance data in Figure S13). Data are shown as mean of two independent measurements. B) Spearman correlation between anti-PEG IgG or IgM titres and C1q binding absorbance of plasma collected before vaccination (Pre-Vax) and post-boost of the mRNA-1273 vaccination (*n* = 40 from 20 donors, donor 56−75, two time points).

Many PEG-based therapeutics are delivered intravenously to promote biodistribution throughout the body and anti-PEG antibodies can interfere with this process. Intramuscular vaccination, however, exerts its effect primarily via the vaccine traversing lymphatics and stimulating immunity in the draining lymph node. This process may be less impacted by systemic anti-PEG antibodies. The impact of PEG-specific antibodies on human LNP vaccination was analyzed by quantifying SARS-CoV-2-specific neutralizing antibody levels 2 to 7 weeks after the second vaccination of BNT162b2 using a live virus neutralization assay, since neutralization responses have emerged as a key correlate of protective immunity.^31^ All 55 BNT162b2 vaccinated subjects generated neutralizing antibody responses to SARS-CoV-2 after the second vaccine (median 346, range 103−1848 ID_50_). There was no correlation between the absolute levels or change of PEG-specific IgG or IgM antibodies before or after vaccination and the neutralizing antibody titres (Figure S14). This suggests PEG antibodies induced by 2 doses of BNT162b2 vaccination do not interfere with vaccine immunogenicity.

There has been speculation that anti-PEG antibody responses may be involved in reactogenicity and anaphylaxis associated with COVID-19 lipid mRNA nanoparticle vaccination.^32-36^ Anaphylaxis and other manifestations of hypersensitivity are rare adverse events after SARS-CoV-2 LNP mRNA vaccination,^11^ while systemic and local reactogenicity after the mRNA-1273 and BNT162b2 vaccination is common.^37^ We assessed self-reported local and systemic reactions after 2 doses of the BNT162b2 and mRNA-1273 vaccination from a subset of 63 of the 75 vaccinees where reactogenicity data was available. Local (at the injection site), systemic (constiutional symptoms) and total reactogenicity was scored as the sum of self-reported local, systemic and total adverse events, respectively (Tables S5, S6). Vaccine reactions were generally mild and no subjects experienced anaphylaxis or myocarditis in our cohort. We found the increase in levels of anti-PEG IgG and IgM correlated with a higher rate of systemic (and total) reactogenicity post-boost of vaccination (Figure 8A,B). In contrast, no association between local reactogenicity and change in anti-PEG antibody was observed (Figure 8A,B). Our study suggests that boosting of anti-PEG antibodies by mRNA LNP vaccination is associated with higher systemic reactogenicity of SARS-CoV-2 LNP mRNA vaccination. Larger cohort studies and animal studies are required to confirm the causal relationship between reactogenicity and change in anti-PEG antibodies.

**Figure 8.**
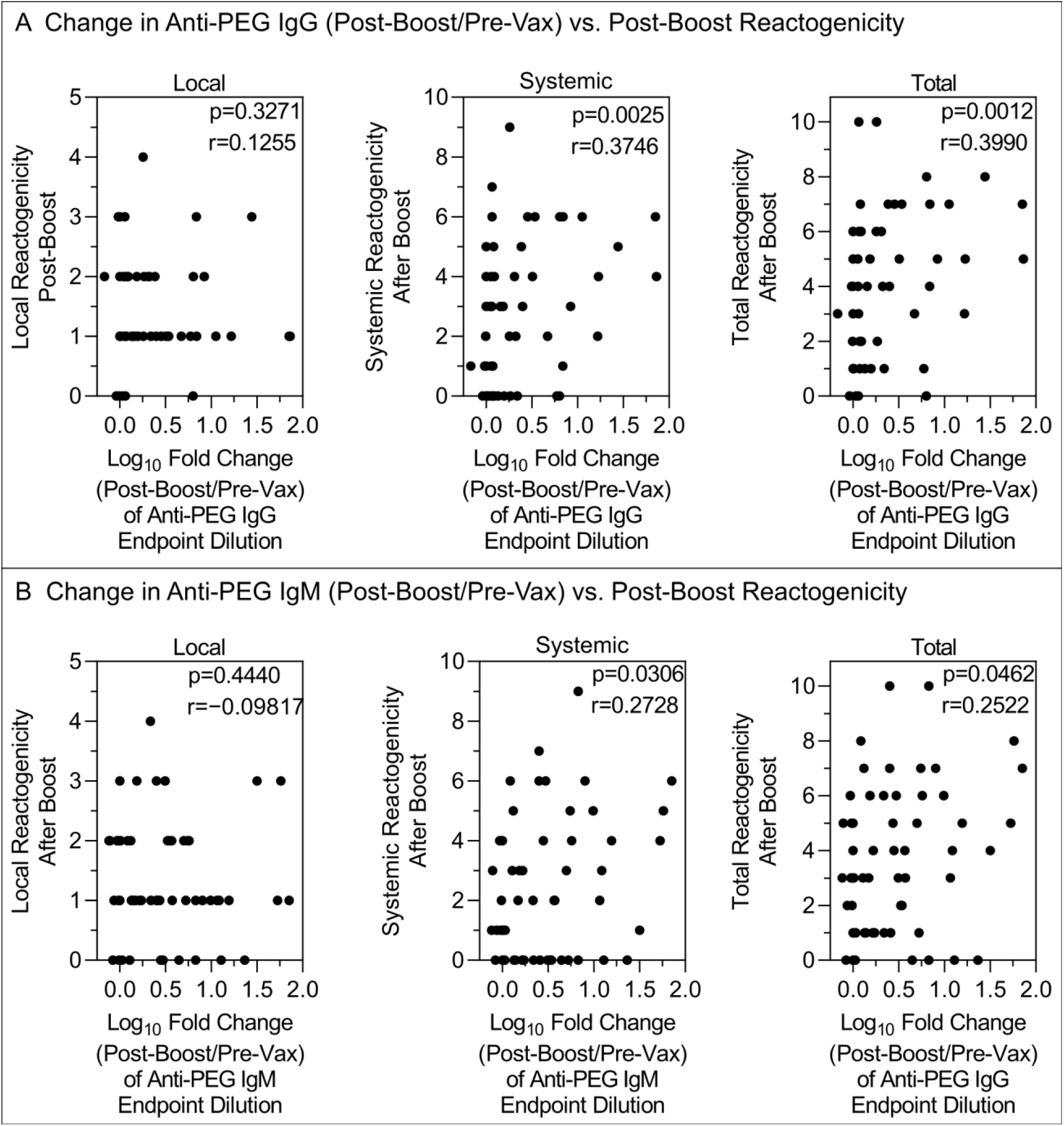
The impact of change in PEG specific antibody on reactogenicity after 2 doses of SARS-CoV-2 LNP mRNA (BNT162b2 or mRNA-1273) vaccination. A, B) Spearman correlation between the fold change (log_10_) of anti-PEG IgG or IgM titres (Post-Boost/Pre-Vax) and post-boost reactogenicity (*n* = 63, including 44 BNT162b2 and 19 mRNA-1273 vaccinees that have reactogenicity reports available, see raw data in Tables S5 and S6).

## CONCLUSIONS

We investigated serial plasma samples from 130 adults and observed that PEG-specific IgG and IgM antibodies were boosted by mRNA-1273 vaccination and to a lesser extent by BNT162b2 vaccination. Although pre-existing levels of anti-PEG antibody were sex- and age-dependent, the boost level of anti-PEG antibody following vaccination was not affected by sex or age. The impact of vaccine-induced anti-PEG antibody on nanoparticle−immune cell interactions was studied using an *ex vivo* human blood assay. We found significant increases in blood cell association with clinically relevant PEGylated LNPs mediated by the anti-PEG antibodies in human plasma after mRNA-1273 vaccination. The PEG-specific antibodies induced by mRNA-1273 vaccination were also able to bind the complement protein C1q in a proportion of subjects, which could potentially result in complement opsonization of nanoparticles. Our study suggests it will be important to monitor the impact of mRNA LNP vaccination on the biology of other PEGylated medicines. We further studied the clinical impact of anti-PEG antibodies induced by the vaccination. PEG-specific antibodies present prior to or following BNT162b2 vaccination did not negatively impact the neutralization immune response to 2 doses of the vaccine. However, we found the increase in anti-PEG antibodies correlated with higher systemic reactogenicity after 2 doses of SARS-CoV-2 LNP mRNA (BNT162b2 and mRNA-1273) vaccination. Larger and longer studies are needed to analyze the longer-term impact of boosting anti-PEG antibodies by LNP mRNA vaccination. Booster doses of SARS-CoV-2 LNP mRNA vaccines, now commonly recommended to enhance immunity^38^ and for emerging variants of SARS-CoV-2,^39^ as well as new mRNA LNP vaccines and therapies for other infectious diseases or cancers^40^ may further increase PEG-specific antibodies. Generating next-generation LNPs with alternatives to PEG^41^ may be useful for overcoming PEG immunogenicity in the future.

## METHODS

### Human Subjects

Seventy-five healthy adults were recruited to donate blood samples before and after receiving the BNT162b2 (*n* = 55) or mRNA-1273 (*n* = 20) LNP mRNA vaccine intramuscularly. Healthy unvaccinated subjects (*n* = 15) followed over time were recruited prior to the COVID-19 pandemic with 2 plasma samples stored at approximately a 6-month interval. In addition, COVID-19 infected subjects (*n* = 40) followed over time with 2 plasma samples stored at approximately a 6-month interval were also studied as unvaccinated controls. Subject details and timing of sampling are shown in Tables S1−S3. All subjects provided written informed consent and the studies were approved by the University of Melbourne human research and ethics committee (approvals 2056689, 21198153983 and 2021-21626-15756-1) and the Royal Melbourne Hospital human research and ethics committee (approvals 6309/MH-2020, 2013.290 and 68355/MH-2020). For all participants, whole blood was collected with sodium heparin anticoagulant. Plasma was collected, aliquoted, and stored at –80 °C. Adverse Events were collected from subjects after vaccination using a self-reported questionnaire (Tables S5 and S6). Reactogenicity scores were calculated by the summing the number of the positive adverse events.

### ELISA to Detect Anti-PEG Antibodies in Human Plasma

The ELISA was conducted based on a modified previously developed method.^3^ The 8-arm PEG-NH_2_ (40 kDa, 200 µg mL^-1^, JenKem Technology, USA) in PBS was coated onto MaxiSorp 96-well plates (Nunc, Denmark) for 18 h at 4 °C, followed by washing with PBS four times. Plates were blocked with 5% (w/v) skim milk powder in PBS for 5 h when detecting anti-PEG IgG or 22 h when detecting anti-PEG IgM, followed by adding serially diluted human plasma in 5% skim milk in duplicate for 1 h at 22 °C. Plates were washed with 0.1% 3-[(3-cholamidopropyl)-dimethylammonio]-1-propanesulfonate (CHAPS, Sigma-Aldrich, USA)/PBS buffer twice and PBS four times prior to addition of an horseradish peroxidase (HRP)-conjugated anti-human IgG (Dako Agilent, USA) at 1:20,000 dilution or HRP-conjugated anti-human IgM (Jackson ImmunoResearch Laboratories, USA) at 1:10,000 for 1 h at 22 °C. Plates were washed as above and then developed using 3,3’,5,5’-Tetramethylbenzidine (TMB) liquid substrate (Sigma-Aldrich, USA). Reaction was stopped with 0.16M H_2_SO_4_ and read at 450 nm. Endpoint titres were calculated as the reciprocal plasma dilution giving signal 2× background using a fitted curve (4-parameter log regression) and reported as a mean of duplicates. Background was detected by adding the diluted plasma samples (at 1:10 dilution in 5% skim milk) to the non-PEG-coated wells, followed by the same ELISA procedure.

### ELISA to Detect Plasma Antibody Response to the Synthetic Lipid Components of BNT162b2 Vaccine

Lipid components 2-[(polyethylene glycol)-2000]-N,N ditetradecylacetamide (ALC-0159, Sinopeg, China), (4-hydroxybutyl) azanediyl)bis (hexane-6,1-diyl)bis(2-hexyldecanoate) (ALC-0315, Sinopeg, China), and 1,2-distearoyl-sn-glycero-3-phosphocholine (DSPC, Avanti Polar Lipids, USA) in 100% ethanol were coated onto PolySorp 96-well plates (Nunc, Denmark) at 100 µg mL^-1^ for 20 h at 37 °C, followed by washing with PBS four times. Plates were blocked with 5% (w/v) skim milk powder in PBS for 5 h, followed by adding serially diluted human plasma in 5% skim milk in duplicate for 1 h at 22 °C. The humanized monoclonal anti-PEG IgG (Hu-6.3-IgG, Institute of Biomedical Sciences at Academia Sinica, Taiwan) was serially diluted in 5% skim milk in duplicate and added to PolySorp 96-well plates for 1 h at 22 °C. Plates were then washed and treated in the same way as the section above.

### ELISA to Detect Anti-PEG Antibody-Mediated C1q Binding

Complement C1q protein were purchased from MP Biomedicals (USA) and were conjugated with biotin using Sulfo-NHS-LC-LC-Biotin (Thermo Scientific, USA) according to the manufacturer’s instruction. The 8-arm PEG-NH_2_ (40 kDa, 200 µg mL^-1^, JenKem Technology, USA) in PBS was coated onto MaxiSorp 96-well plates (Nunc, Denmark) for 18 h at 4 °C, followed by washing with PBS four times. Plates were blocked with 5% (w/v) bovine serum albumin (BSA) in PBS for 18 h, followed by adding diluted human plasma at 1:5 in 1% BSA in duplicate for 1 h at 22 °C. Biotinylated C1q and HRP-conjugated streptavidin (Thermo Scientific, USA) were mixed at a molar ratio of 4:1 for 30 mins at 22 °C. After plasma incubation, plates were washed with 0.1% CHAPS/PBS buffer twice and PBS four times prior to addition of the pre-mixed biotinylated C1q and HRP-conjugated streptavidin at 2 ug mL^-1^ of C1q for 2 h at 22 °C. Plates were washed as above and then developed using TMB liquid substrate (Sigma-Aldrich, USA). Reaction was stopped with 0.16M H_2_SO_4_ and read at 450 nm. Non-PEG-precoated wells were treated the same as PEG-precoated wells and used as background for absorbance subtraction.

### Particle Preparation

Alexa Fluor 647 (AF647)-labelled PEGylated mesoporous silica (PEG-MS) nanoparticles were prepared using a previously published method.^19^ Dioctadecyl-3,3,3,3-Tetramethylindodicarbocyanine (DiD)-labelled Doxoves, PEGylated doxorubicin-encapsulated liposomes that contain the same lipid composition and drug/lipid ratio as clinically used liposomal doxorubicin agent (Doxil), in addition to 0.2 mol% DiD, were purchased from FormuMax Scientific Inc, USA. DiD-labelled PEGylated LNPs were formulated with (6Z,9Z,28Z,31Z)-heptatriaconta-6,9,28,31-tetraen-19-yl 4-(dimethylamino)butanoate (DLin-MC3-DMA, DC Chemical, China), DSPC (Avanti Polar Lipids, USA), cholesterol (Sigma-Aldrich, USA), and 1,2-dimyristoyl-rac-glycero-3-methoxypolyethylene glycol-2000 (PEG2000-DMG, Avanti Polar Lipids, USA) with the same molar composition of lipids used in the US FDA-approved Onpattro (Patisiran) formulation, in addition to 0.1 mol% DiD (Thermo Fisher Scientific, USA), using the NanoAssemblr platform (Precision NanoSystems, Canada). Particles were loaded with a non-immunogenic nucleic acid cargo (firefly luciferase plasmid DNA, PlasmidFactory GmbH & Co. KG, Germany) in order to regulate lipid packing and particle size.

### Particle Characterization

Transmission electron microscopy (TEM) images were acquired using an FEI Tecnai TF20 instrument at an operation voltage of 120 kV under liquid nitrogen cooling. PEG-MS particle suspensions were dropped and air-dried on Formvar carbon-coated copper grids (plasma-treated). Cryogenic TEM (Cryo-TEM) images were acquired using a FEI Tecnai Spirit instrument at an operation voltage of 120 kV under liquid nitrogen cooling under low dose conditions. Doxil and LNP suspensions were applied to glow-discharged lacey carbon grids and vitrified using a Vitrobot (FEI) system. Vitrified sample grids were stored in liquid nitrogen prior to imaging. Dynamic light scattering (DLS) analysis of the particles was performed on a Zetasizer Nano-ZS (Malvern Instruments, UK) instrument. Zeta-potential measurements of the particles were performed at pH 7.4 in phosphate buffer (5 mM) using a Zetasizer Nano-ZS (Malvern Instruments, UK). The mass of the PEG-MS particles was determined by weighing the freeze-dried particles. The concentration of Doxil (based on lipid) was provided by the manufacturer. The concentration of encapsulated pDNA in Onpattro LNPs (97 μg/ml) was determined using the Quant-iT™ PicoGreen™ dsDNA Assay kit; encapsulation efficiency of LNPs utilized in this study was >96%.

### Blood Assay to Determine Nanoparticle Association with Human Immune Cells

The human blood assay was conducted based on a modified previously developed method.^16^ The fresh human blood was collected from a healthy volunteer (donor 16) into sodium heparin vacuettes (Greiner Bio-One). The blood was centrifuged (900*g*, 15 min, no brake), followed by removing plasma in the top layer. The remaining blood cells were further washed with serum-free RPMI 1640 medium for five times to remove any residual plasma proteins. The absence of plasma proteins was confirmed by the lack of absorbance of the supernatant at 280 nm (Nanodrop 2000, Thermo Fisher Scientific, USA). Washed blood cells were finally dispersed in serum-free RPMI 1640 medium and counted with a CELL-DYN Emerald analyzer. Nanoparticles (3.2 µg PEG-MS, 3.2 µg Doxil, or 1 µg LNP) were preincubated in plasma (100 µL) from different subjects (donor 1−18 from BNT162b2 vaccinee cohort or donor 56−75 from mRNA-1273 vaccinee cohort, collected before vaccination vs. post-boost vaccination) at 37 °C for 1 h. Individual plasma from different subjects at each time point (pre-vax vs. post-boost) were used to coat onto nanoparticles, forming a personalized protein corona. The nanoparticles (0.2 µg) with a personalized plasma protein corona were subsequently added to washed blood cells (5 × 10^5^) in serum-free RMPI 1640 medium (100 µL) at a particle concentration of 2 µg mL^-1^ for 1 h at 37 °C. After incubation, the red blood cells were lysed using Pharm Lyse buffer (4 mL) and removed by centrifugation (500 g, 5 min). After washing twice with flow cytometry staining (FACS) wash buffer (PBS containing 2 mM EDTA and 0.5% w/v BSA), the cells were phenotyped at 4 °C for 1 h using an antibody cocktail consisting of CD3 AF700 (SP34-2, BD), CD14 APC-H7 (MΦP9, BD), CD66b BV421 (G10F5, BD), CD45 V500 (HI30, BD), CD56 PE (B159, BD), lineage-1 (Lin-1) cocktail FITC (BD), HLA-DR PerCP-Cy5.5 (G46-6, BD), and CD19 BV650 (HIB19, BioLegend) in titrated concentrations. The cells were subsequently washed twice with FACS wash buffer (4 mL, 500 g, 7 min) to remove unbound antibodies and fixed with 1% w/v formaldehyde in PBS and directly analyzed by flow cytometry (LSRFortessa, BD Bioscience). The data were processed using FlowJo V10. Cell association (%) refers to the proportion of each cell type with positive fluorescence, above background, stemming from fluorescence-labelled nanoparticles (gating strategy shown Figure S5). Data was reported as a mean of three independent experiments (using the same batch of plasma from each donor), with at least 150,000 leukocytes analyzed for each experimental condition studied. Cell association data of T cells, NK cells, and dendritic cells were uniformly low (data not shown) and thus excluded from further data analysis.

### SARS-CoV-2 Live Virus Neutralization Assay

A SARS-CoV-2 isolate (CoV/Australia/VIC/01/2020) was passaged in Vero cells and stored at -80ºC. 96-well flat bottom plates were seeded with Vero cells (20,000 cells per well in 100µl). The next day, Vero cells were washed once with 200 µl serum-free DMEM prior to the addition of 150µl infection media (serum-free DMEM with 1.33 µg/ml TPCK trypsin). Eight 2.5-fold serial dilutions of heat-inactivated sera (from 1:20 to 1:12,207) were incubated with SARS-CoV-2 virus at 2000 TCID_50_/ml at 37ºC for 1 hour. Next, sera-virus mixtures (50µl) were added to Vero cells in duplicate and incubated at 37 ºC for 48 hours. ‘Cells only’ and ‘virus+cells’ controls were included to represent 0% and 100% infectivity respectively. After 48 hours, all cell culture media were carefully removed from wells and 200 µl of 4% formaldehyde was added to fix the cells for 30 mins at room temperature. The plates were then dunked in a 1% formaldehyde bath for 30 minutes to inactivate any residual virus prior to removal from the BSL3 facility. Cells were washed once in PBS and then permeabilized with 150 µl of 0.1% Triton-X for 15 minutes. Following one wash in PBS, wells were blocked with 200µl of blocking solution (4% BSA with 0.1% Tween-20) for 1 hour. After three washes in PBST (PBS with 0.05% Tween-20), wells were added with 100µl of rabbit polyclonal anti-SARS-CoV N antibody (Rockland, #200-401-A50) at a 1:8000 dilution in dilution buffer (PBS with 0.2% Tween-20, 0.1% BSA and 0.5% NP-40) for 1 hour. Plates were then washed six times in PBST and added with 100µl of goat anti-rabbit IgG (Abcam, #ab6721) at a 1:8000 dilution for 1 hour. After six washes in PBST, plates were developed with TMB and stopped with 0.15M H_2_SO_4_. OD values read at 450 nm were then averaged for duplicate samples and used to calculate %neutralization with the following formula: (‘Virus + cells’ – ‘sample’) ÷ (‘Virus + cells’ – ‘Cells only’) × 100. Inhibitory dilution 50 (ID_50_) values were determined using four-parameter nonlinear regression in GraphPad Prism with curve fits constrained to have a minimum of 0% and maximum of 100% neutralization.

### Statistical Analyses

Pair-comparison in Figures 1A–C, 3A, 6A, S4B (between 1-Month Post-Boost and 3-Month post-Boost), S10A, and S12A was derived by Wilcoxon’s matched-pairs signed rank test using Prism 9.0 (GraphPad). Cross-comparison between the three cohorts in Figure 1D were derived by nonparametric Kruskal-Wallis test with Dunn’s multiple comparisons test using Prism 9.0 (GraphPad). Statistical significance in Figure 2A,C was assessed by Mann-Whitney *U* test using Prism 9.0 (GraphPad). Longitudinal comparison in Figure S4 was derived by nonparametric Friedman’s test with Dunn’s multiple comparisons test in Prism 9.0 (GraphPad). Correlation analysis in Figures 2B,D, 3B, 5, 6B, 7B, 8, S9, S10B, S11, S12B, and S14 were assessed using nonparametric Spearman correlation tests in Prism 9.0 (GraphPad).

**Scheme 1.**
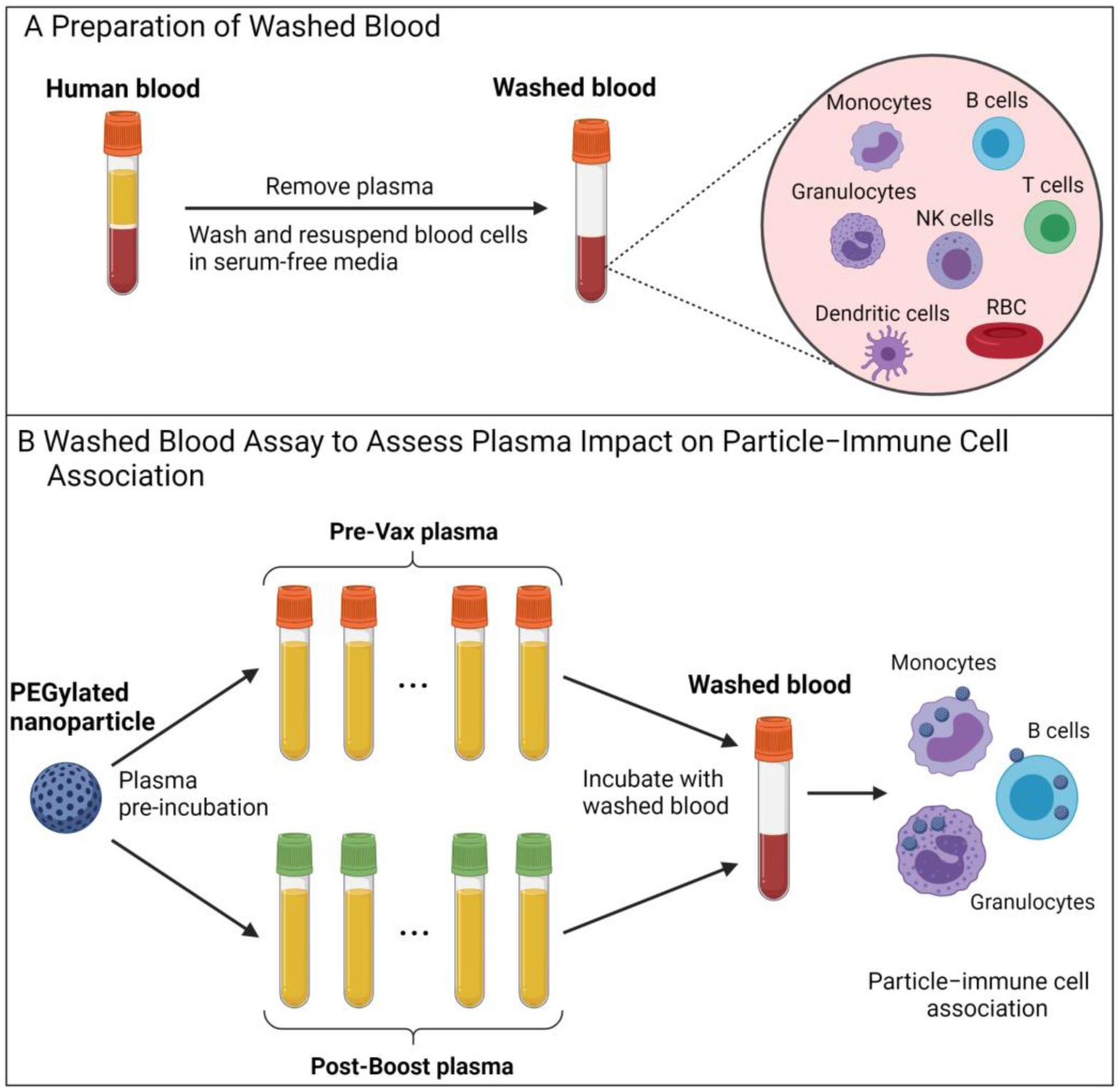
Schematic illustration of human blood assay to assess the impact of plasma on PEGylated nanoparticle association with human immune cells.^a^ ^a^Fresh human blood from a healthy donor was washed by centrifugation with serum-free media multiple times to completely remove plasma. PEGylated nanoparticles were pre-incubated with plasma from multiple donors before vaccination (Pre-Vax) and post-boost of BNT162b2 or mRNA-1273 vaccination, and then incubated with washed blood in serum-free media for 1 h at 37 °C, followed by phenotyping cells with antibody cocktails and analysis by flow cytometry. Created with BioRender.com.

## Data Availability

All data produced in the present work are contained in the manuscript and supporting information

## ASSOCIATED CONTENT

### Supporting Information

Time course of anti-PEG IgG and IgM levels of three cohort: BNT162b2, mRNA-1273, and unvaccinated control, gating strategy used to identify white blood cell populations, LNP, Doxil and PEG-MS cell association data, the impact of anti-PEG IgG or IgM on nanoparticle association with human blood immune cells, anti-PEG antibody-induced C1q binding, neutralizing antibody responses, BNT162b2 vaccinee information, mRNA-1273 vaccinee information, and unvaccinated control blood donor information, reported reactogenicity after boost of BNT162b2 and mRNA-1273 vaccination.

## AUTHOR INFORMATION

### Author Contributions

Y.J. and S.J.K. conceived, designed, and supervised the study and drafted the manuscript. Y.J., W.S.L., E.H.P., H.G.K., S.L., K.J.S., and J.A.J. performed experiments and provided technical advice. K.M.W., K.S., T.H.O.N., L.C.R., L.F.A., K.B., D.A.W., K.K., S.M., A.K.W., J.A.J., and S.J.K. recruited subjects and processed their blood samples. N.P.T., M.P., A.W.C., and F.C. provided intellectual input and reagents. All authors approved the final version of the manuscript.

### Notes

The authors declare no competing financial interest.

## ACKNOWLEDGMENTS

This work was supported by the Australian National Health and Medical Research Council (NHMRC) fellowship or leadership awards to S.J.K., K.K., J.A.J., A.K.W., F.C., A.W.C., D.A.W., K.S. and T.H.O.N.; RMIT Vice Chancellor’s Postdoctoral Fellowship to Y.J.; the Australian Medical Research Future Fund award #2005544 to K.K., S.J.K., J.A.J., A.K.W., A.W.C. and D.A.W.; Australian Research Council awards DP210103114 and DP200100231 to F.C., N.P.T., S.J.K., Y.J. and A.K.W., and NHMRC award 1149990 to S.J.K. and F.C.. The Melbourne WHO Collaborating Centre for Reference and Research on Influenza is supported by the Australian Government Department of Health. We thank all the dedicated participants involved in the studies. We thank T. Amarasena, H. Kent, J. Mitchell, C. Uzoho, B. Scher, D. Rathnayake, H. Ding and L. Randall for expert assistance.

## Notes

### Competing Interest Statement

The authors have declared no competing interest.

### Author Declarations

All subjects provided written informed consent and the studies were approved by the University of Melbourne human research and ethics committee (approvals 2056689, 21198153983 and 2021-21626-15756-1) and the Royal Melbourne Hospital human research and ethics committee (approvals 6309/MH-2020, 2013.290 and 68355/MH-2020).

### Summary of Updates

We have included a new vaccine cohort (Moderna mRNA-1273 vaccinee cohort) to study anti-PEG antibodies, a new clinically relevant lipid nanoparticle (Onpattro) to examine PEG antibody-mediated blood immune cell association, a new anti-PEG antibody-mediated complement protein C1q binding assay, and updated reactogenicity data based on the new mRNA-1273 cohort.

