## Supporting Information for "Anti-PEG Antibodies Boosted in Humans by SARS-CoV-2 Lipid Nanoparticle mRNA Vaccine"

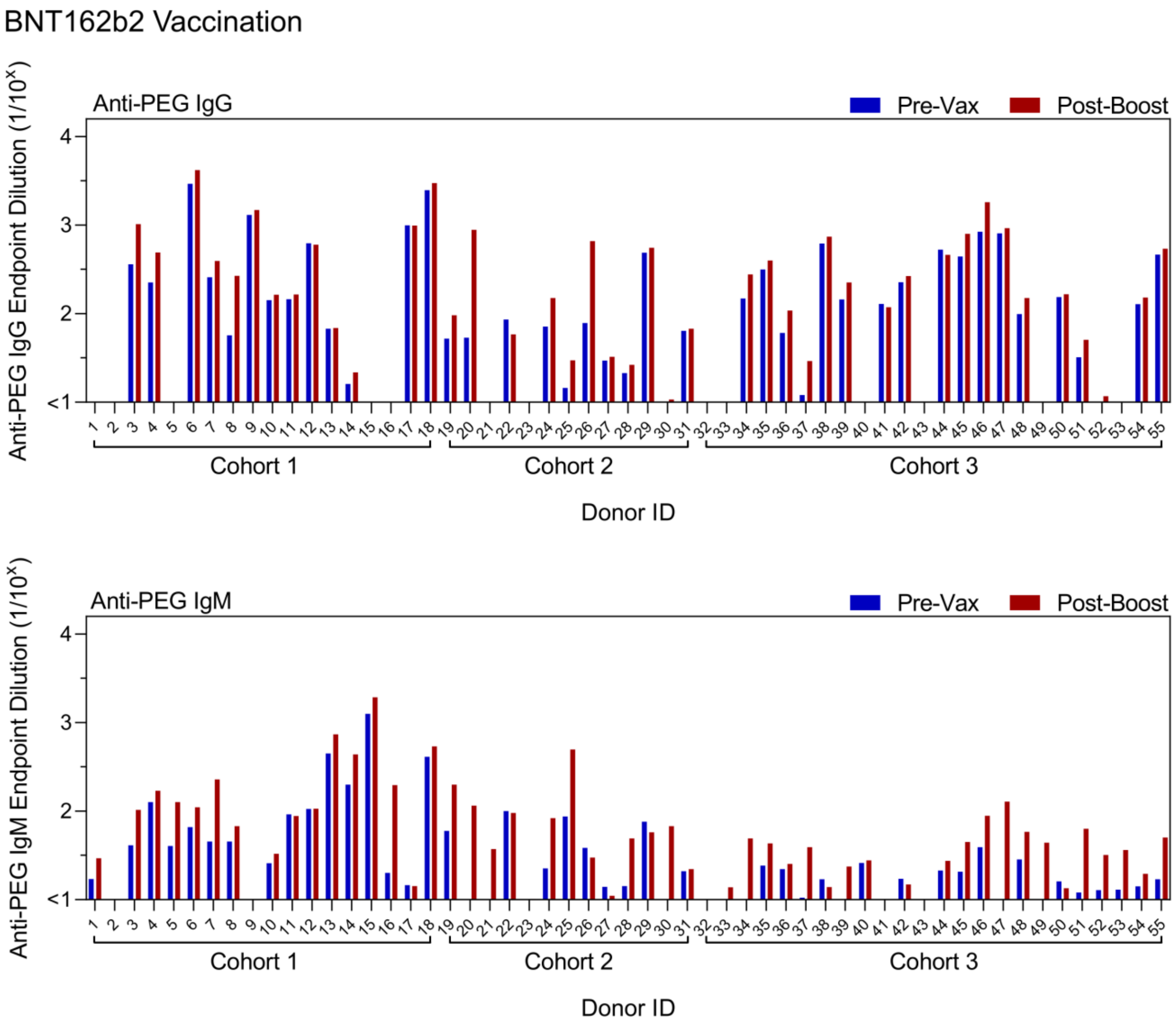


**Figure S1.** Plasma anti-PEG IgG and IgM titres before vaccination (Pre-Vax) and 2 to 7 weeks (mean 27 days, range 12−49) following second dose (Post-Boost) of BNT162b2 vaccination of 55 healthy subjects across 3 separate cohorts. Data are shown as mean of two independent measurements.

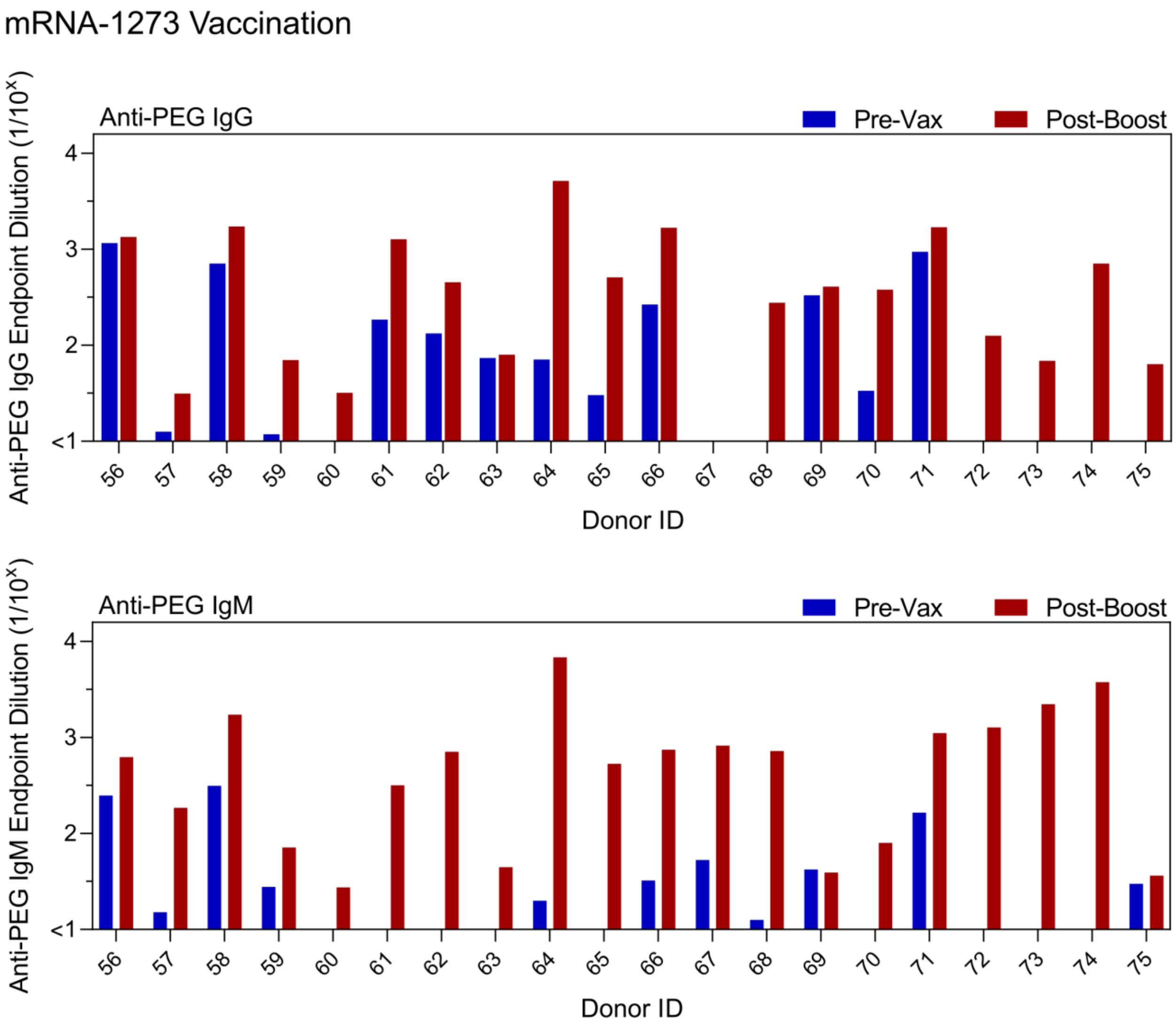


**Figure S2.** Plasma anti-PEG IgG and IgM titres before vaccination (Pre-Vax) and 3 weeks (mean 19 days, range 18−27) following second dose (Post-Boost) of mRNA-1273 vaccination of 20 healthy subjects. Data are shown as mean of two independent measurements.

Unvaccinated Control

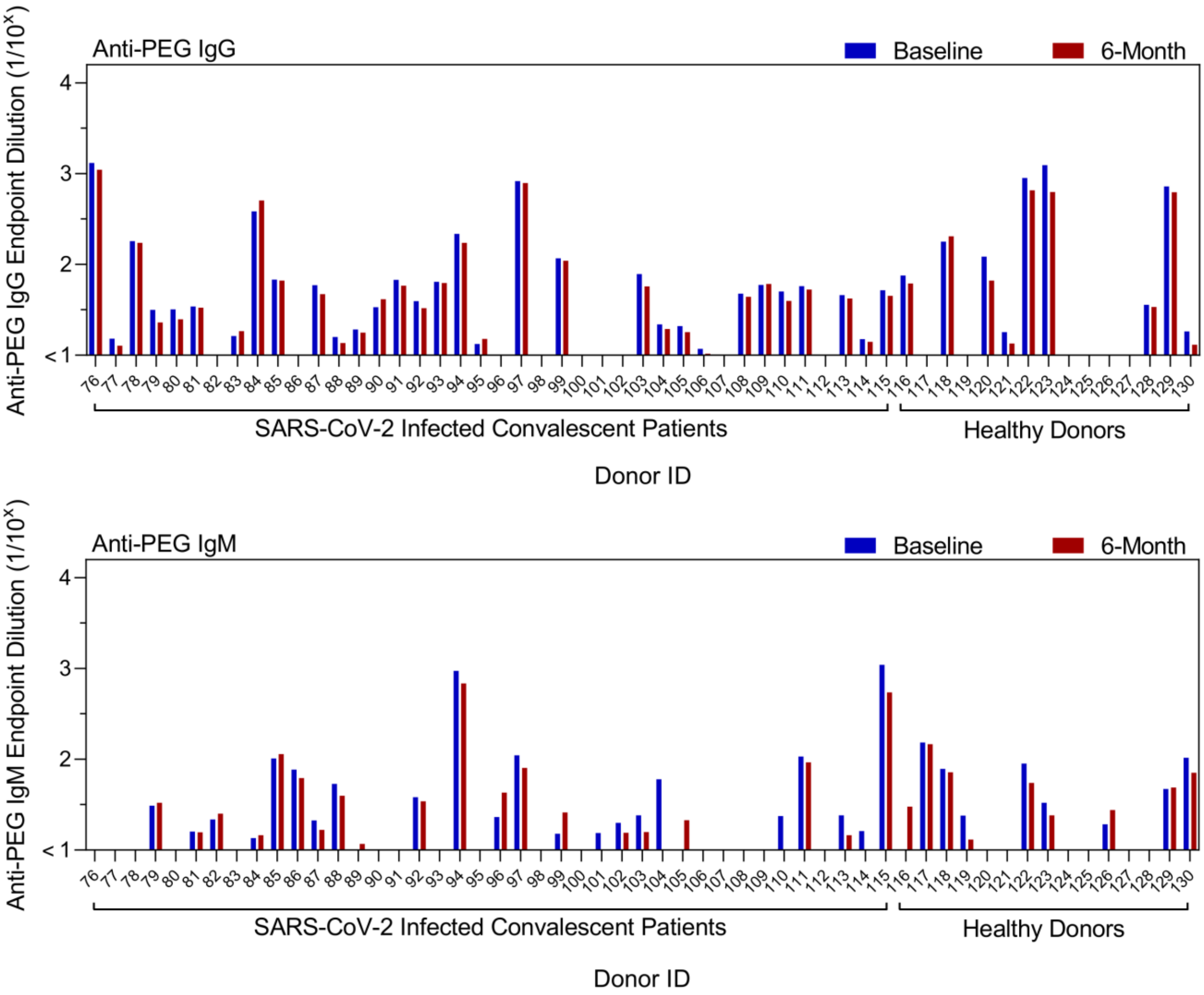


**Figure S3.** Plasma anti-PEG IgG and IgM titres at baseline and after a 6-month (mean 186 days, range 120−253) period of unvaccinated control cohorts (total n=55, including SARS-CoV-2 infected convalescent patients n=40 and uninfected healthy donors n=15). Data are shown as mean of two independent measurements.

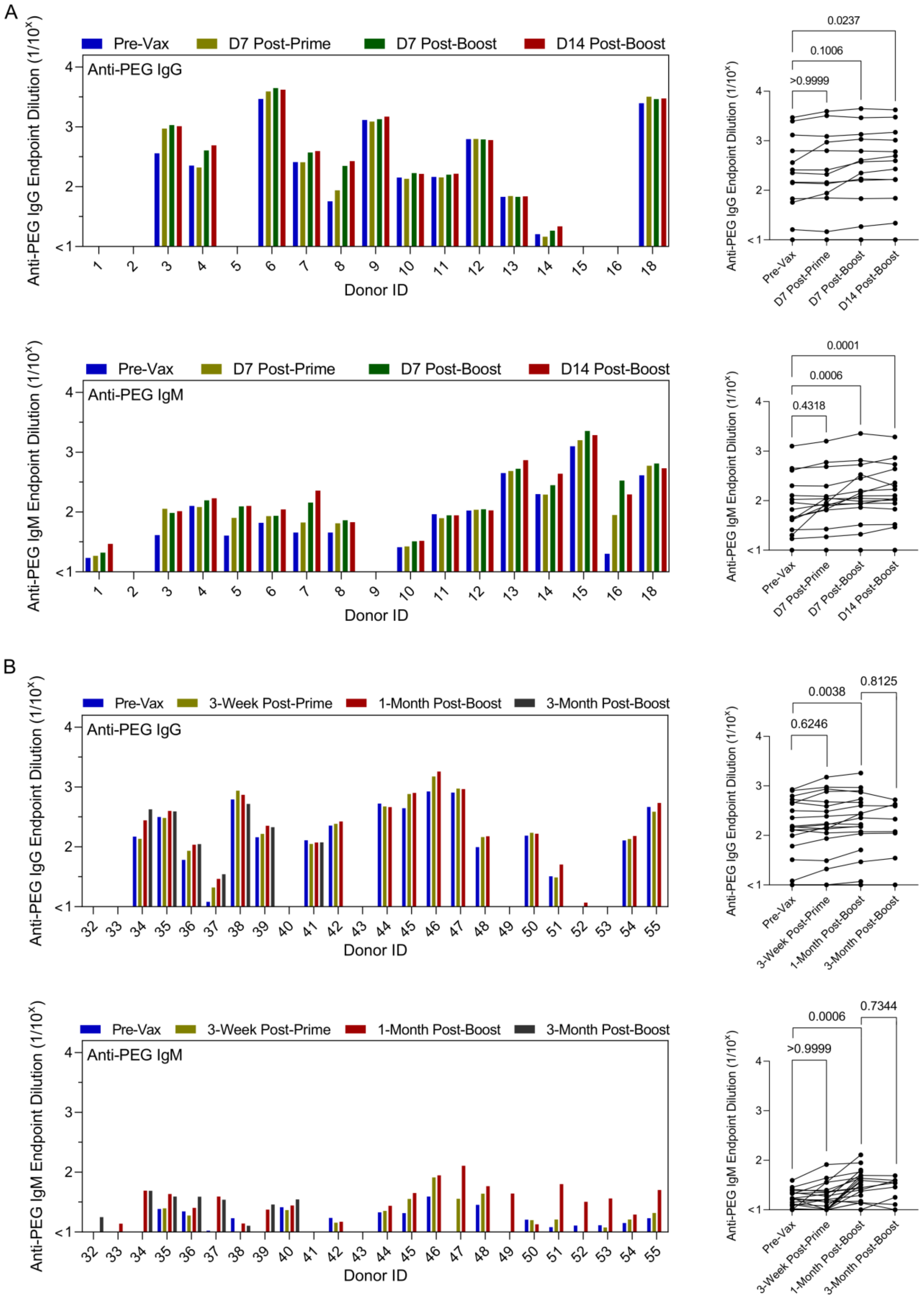
A
Pre-Vax
D7 Post-Prime
D7 Post-Boost
D14 Post-Boost
Anti-PEG IgG
Anti-PEG IgG Endpoint Dilution (1/10^x)
Donor ID
0.0237
0.1006
>0.9999
Anti-PEG IgM
Anti-PEG IgM Endpoint Dilution (1/10^x)
0.0001
0.0006
0.4318
B
Pre-Vax
3-Week Post-Prime
1-Month Post-Boost
3-Month Post-Boost
0.0038
0.8125
0.6246
0.0006
0.7344
>0.9999

**Figure S4**. Time course of PEG antibodies following BNT162b2 vaccination. A) Longitudinal comparison of plasma Anti-PEG IgG and IgM titres in BNT162b2 Cohort 1 subjects (n=17) receiving two doses of the BNT162b2 vaccine at 0−7 days pre-vaccination (Pre-Vax), day 7 post prime, day 7 and 14 post boost. Donor 17's data is excluded due to missing the two middle time points. Data are shown as mean of two independent measurements. p-values were derived by nonparametric Friedman's test with Dunn's multiple comparisons test. B) Longitudinal comparison of plasma Anti-PEG IgG and IgM titres in BNT162b2 Cohort 3 subjects (n=24) receiving two doses of the BNT162b2 vaccine at 0−4 weeks pre-vaccination (Pre-Vax), 3-week (average 22 days, range 14−34) post prime, 1-month (average 33 days, range 22−49) and 3-month (average 85 days, range 74−98) post boost. The 3-month post-boost time points were only collected from 10 subjects (donor 32−41). Data are shown as mean of two independent measurements. p-values comparing between Pre-Vax and 3-Week Post-Prime or 1-Month Post-Boost were derived by nonparametric Friedman's test with Dunn's multiple comparisons test. p-values comparing between 1-Month Post-Boost and 3-Month post-Boost were derived by Wilcoxon's matched-pairs signed rank test.

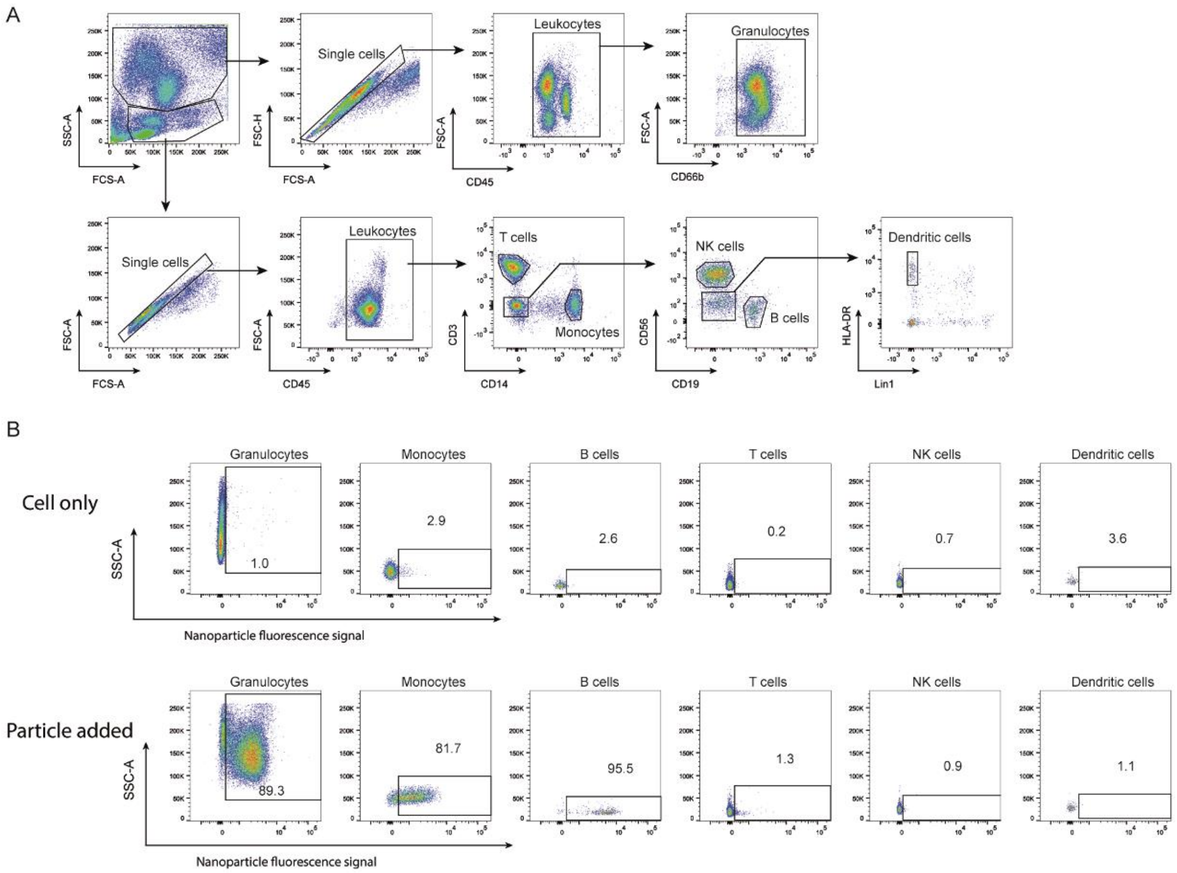


**Figure S5**. Gating strategy used to identify white blood cell populations. A) Flow cytometry gating strategy used to identify different immune cell populations from human blood. Forward and side scattering was used to locate white blood cells, followed by excluding doublets. The following cell types were identified based on the expression of surface markers: CD45+ leukocytes, CD66b+ granulocytes, CD14+ monocytes, CD3+ T cells, CD19+ B cells, CD56+ NK cells, and Lin-HLA-DR+ dendritic cells. B) The percentage of each cell type positive for the fluorescence-labelled nanoparticles was then measured as the cell association (%). The representative gating strategy and cell association percentages for each cell type is shown.

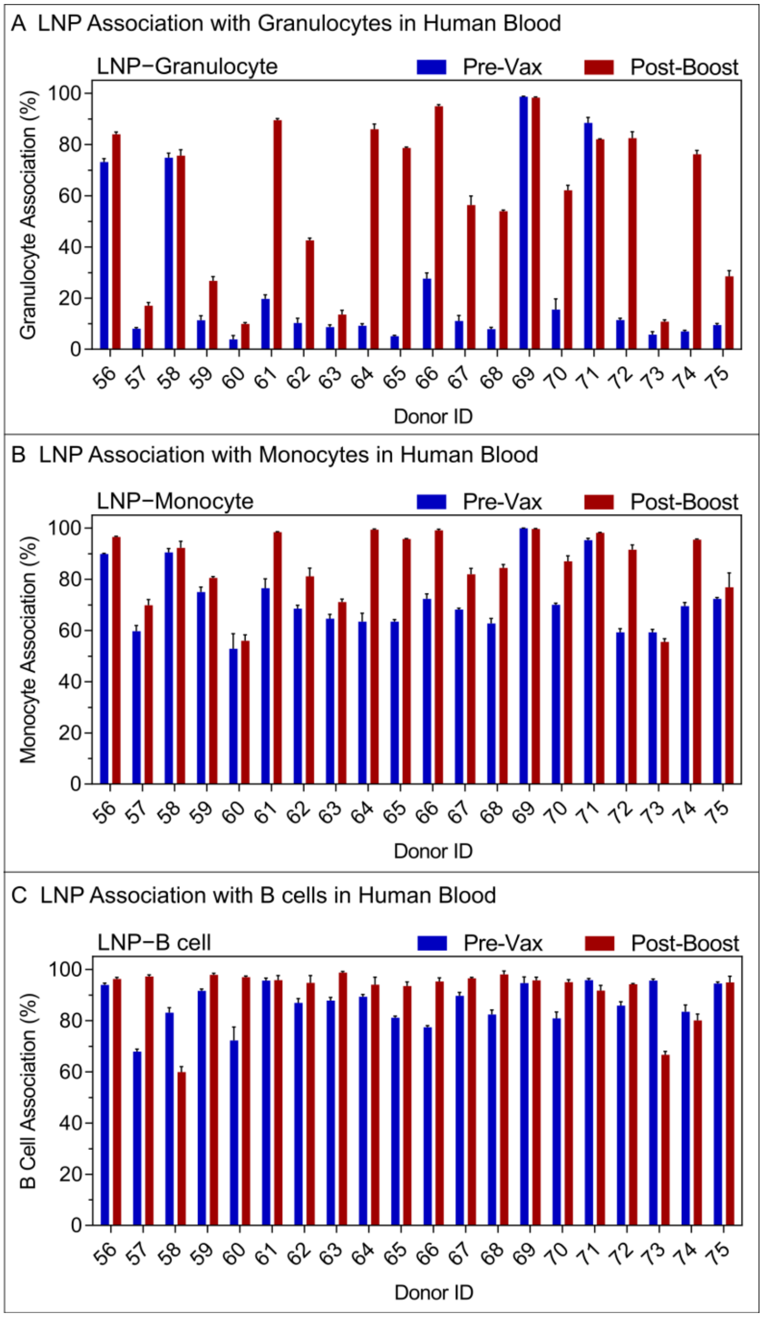


**Figure S6.** (A-C) The impact of plasma collected before vaccination (Pre-Vax) and post-boost of the mRNA-1273 vaccination (n=20, donor 56−75) on Onpattro LNP association with (A) granulocytes, (B) monocytes, or (C) B cells in human blood. Onpattro LNP were preincubated in plasma from donor 56−75 collected Pre-Vax or Post-Boost of the mRNA-1273 vaccination, followed by incubation with washed blood cells from donor 16 in serum-free media. Cell association (%) refers to the proportion of each cell type with positive fluorescence, above background, stemming from fluorescence-labeled particles. Data are shown as the mean ± standard deviation (SD) of three independent experiments (using the same batch of plasma from each donor), with at least 150,000 leukocytes analyzed for each experimental condition studied.

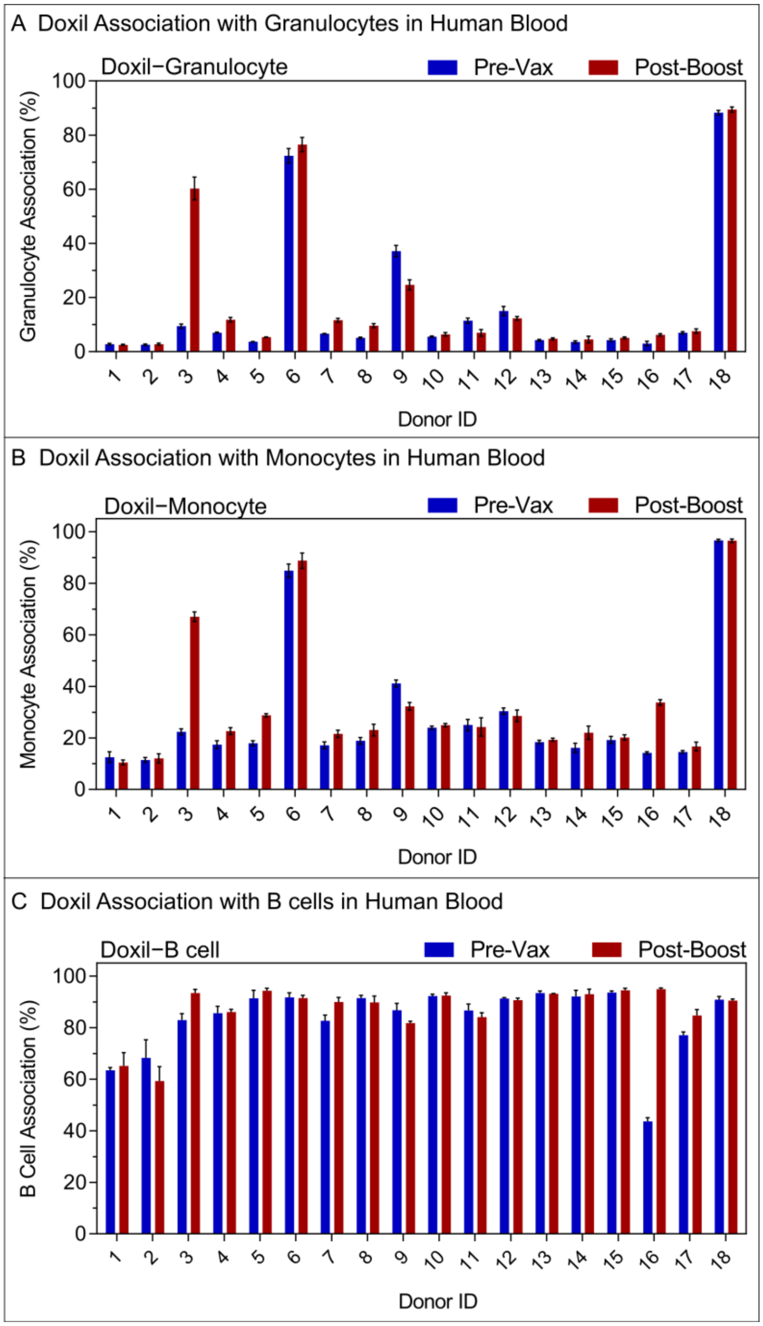


**Figure S7.** (A-C) The impact of plasma collected before vaccination (Pre-Vax) and post-boost of the BNT162b2 vaccination (n=18, donor 1−18) on Doxil association with (A) granulocytes, (B) monocytes, or (C) B cells in human blood. Doxil were preincubated in plasma from donor 1−18 collected Pre-Vax or Post-Boost of the BNT162b2 vaccination, followed by incubation with washed blood cells from donor 16 in serum-free media. Cell association (%) refers to the proportion of each cell type with positive fluorescence, above background, stemming from fluorescence-labeled particles. Data are shown as the mean ± SD of three independent experiments (using the same batch of plasma from each donor), with at least 150,000 leukocytes analyzed for each experimental condition studied.

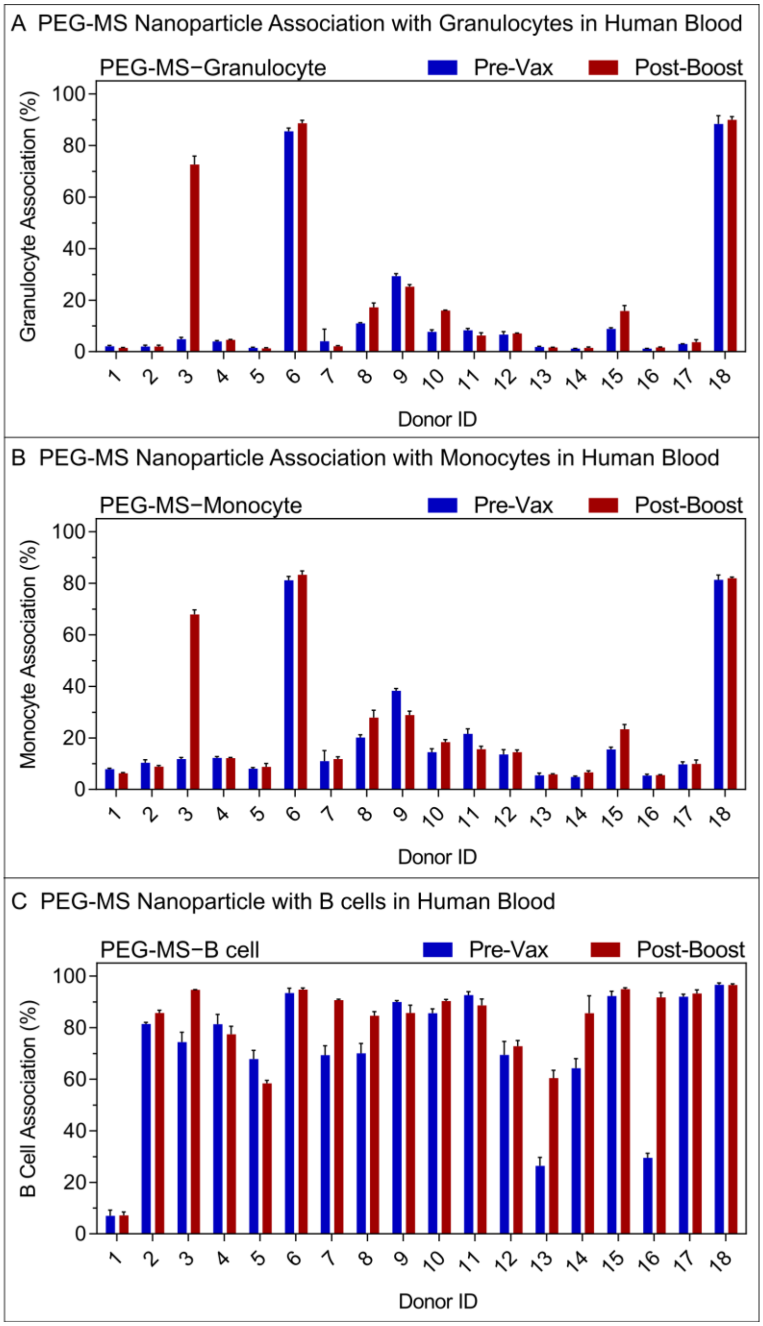


**Figure S8.** (A-C) The impact of plasma collected before vaccination (Pre-Vax) and post-boost of the BNT162b2 vaccination (n=18, donor 1−18) on PEG-MS nanoparticle association with (A) granulocytes, (B) monocytes, or (C) B cells in human blood. PEG-MS nanoparticles were preincubated in plasma from donor 1−18 collected Pre-Vax or Post-Boost of the BNT162b2 vaccination, followed by incubation with washed blood cells from donor 16 in serum-free media.

Cell association (%) refers to the proportion of each cell type with positive fluorescence, above background, stemming from fluorescence-labeled particles. Data are shown as the mean ± SD of three independent experiments (using the same batch of plasma from each donor), with at least 150,000 leukocytes analyzed for each experimental condition studied.

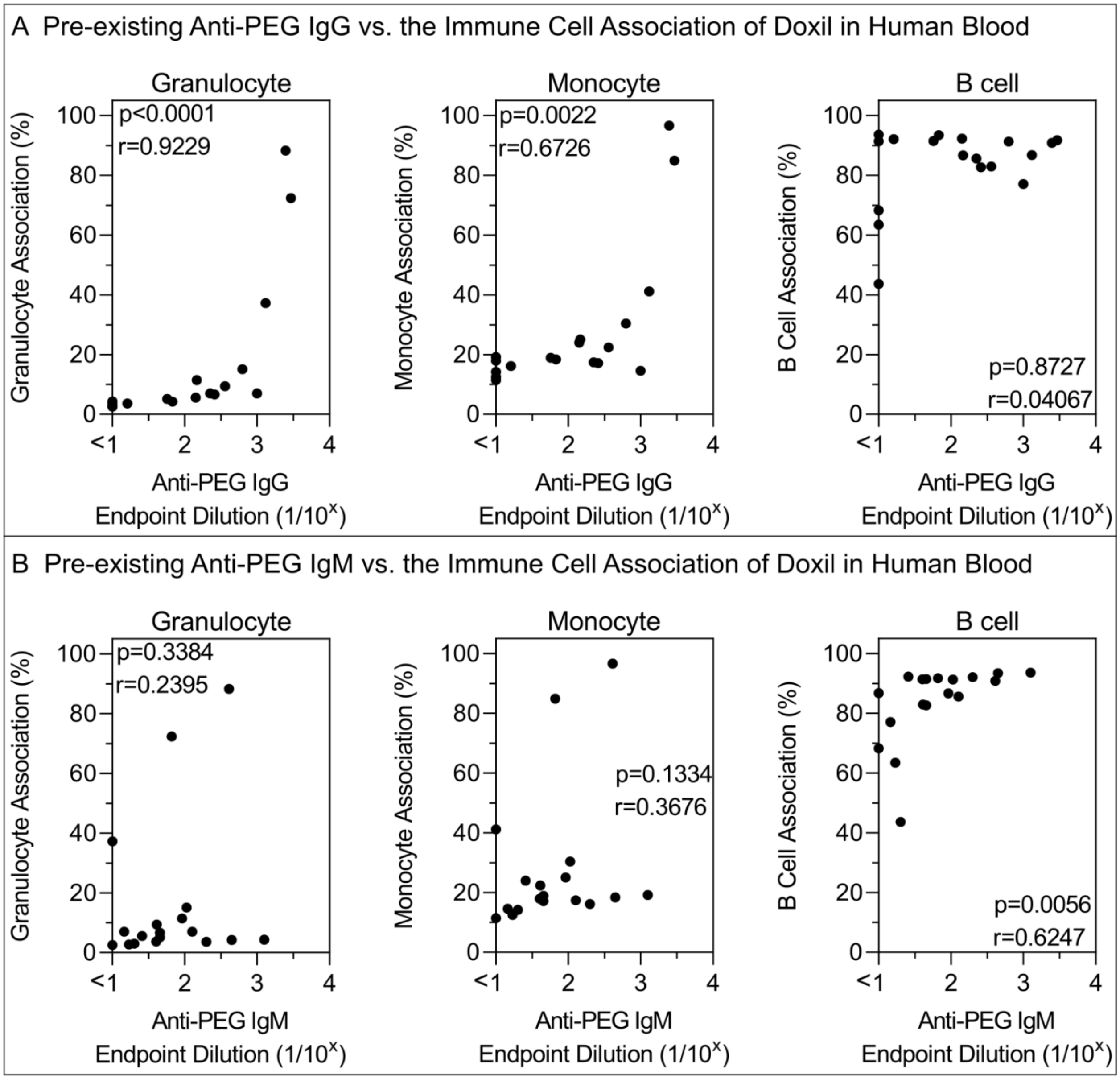


**Figure S9.** Impact of pre-existing anti-PEG antibody levels on the association of Doxil with immune cells in human blood. A,B) Spearman correlation between pre-existing (Pre-Vax) anti-PEG IgG or IgM titres (n=18, donor 1−18 from BNT162b2 vaccinee cohort) and Doxil association with monocytes, granulocytes, or B cells in human blood. Cell association (%) refers to the proportion of each cell type with positive fluorescence, above background, stemming from fluorescence-labeled particles. Cell association (%) data are shown as the mean of three independent experiments (using the same batch of plasma from each donor), with at least 150,000 leukocytes analyzed for each experimental condition studied (see raw data in Figure S7).

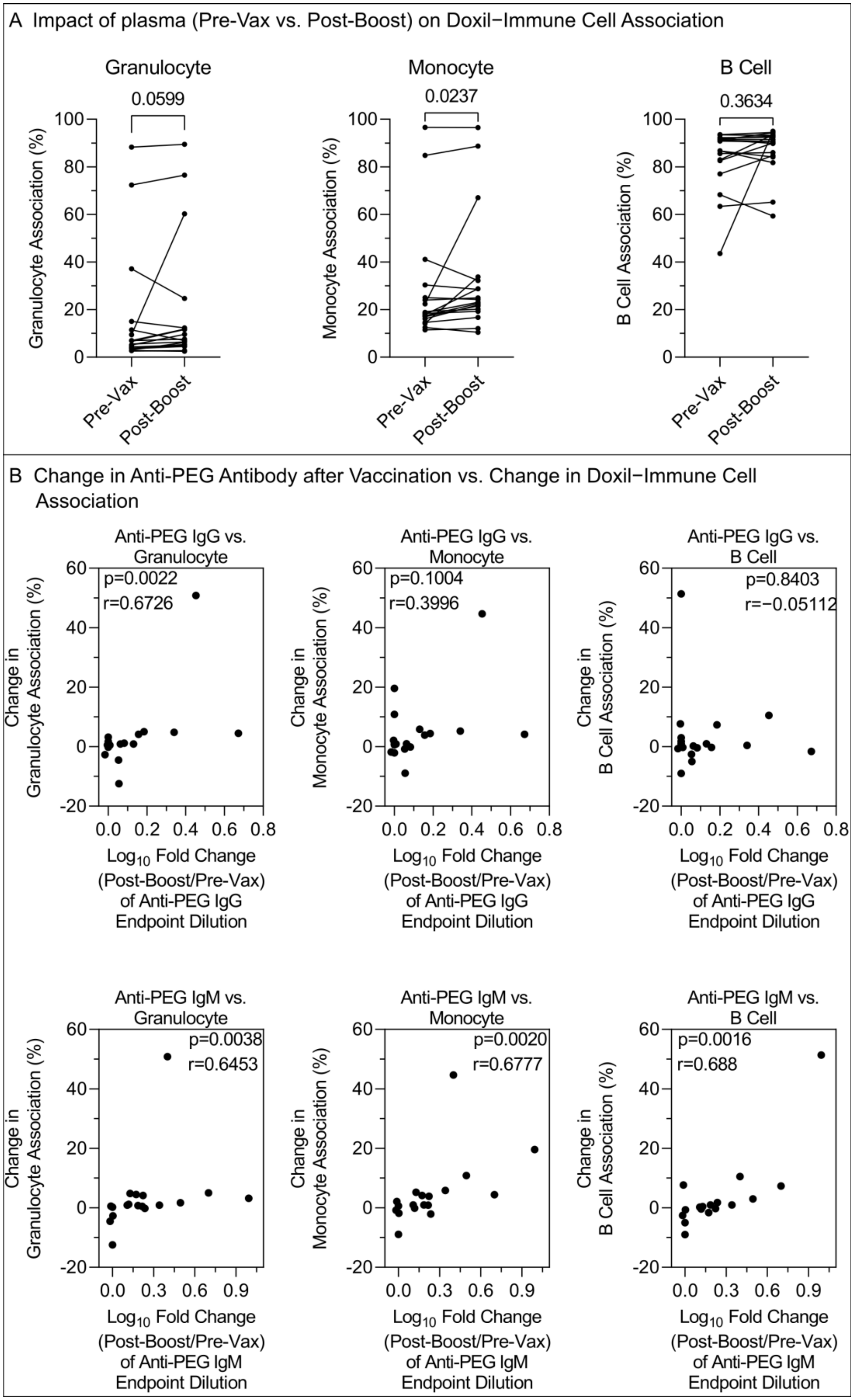
A Impact of plasma (Pre-Vax vs. Post-Boost) on Doxil−Immune Cell Association
Granulocyte
0.0599
Granulocyte Association (%)
Monocyte
0.0237
Monocyte Association (%)
B Cell
0.3634
B Cell Association (%)
Pre-Vax
Post-Boost
B Change in Anti-PEG Antibody after Vaccination vs. Change in Doxil−Immune Cell Association
Anti-PEG IgG vs. Granulocyte
p=0.0022
r=0.6726
Anti-PEG IgG vs. Monocyte
p=0.1004
r=0.3996
Anti-PEG IgG vs. B Cell
p=0.8403
r=−0.05112
Change in Granulocyte Association (%)
Change in Monocyte Association (%)
Change in B Cell Association (%)
Log10 Fold Change (Post-Boost/Pre-Vax) of Anti-PEG IgG Endpoint Dilution
Anti-PEG IgM vs. Granulocyte
p=0.0038
r=0.6453
Anti-PEG IgM vs. Monocyte
p=0.0020
r=0.6777
Anti-PEG IgM vs. B Cell
p=0.0016
r=0.688
Log10 Fold Change (Post-Boost/Pre-Vax) of Anti-PEG IgM Endpoint Dilution

**Figure S10.** Impact of change in anti-PEG antibody after BNT162b2 vaccination on the association of Doxil with immune cells in human blood. A) Comparing the impact of plasma collected before vaccination (Pre-Vax) and post-boost of the BNT162b2 vaccination (n=18, donor 1−18) on Doxil association with monocytes, granulocytes, or B cells in human blood. p-values were derived by Wilcoxon's matched-pairs signed rank test. B) Spearman correlation between the fold change ($log_{10}$) of anti-PEG IgG or IgM titres (Post-Boost/Pre-Vax) after 2 doses of BNT162b2 vaccination (n=18, donor 1−18) and change in Doxil association with monocytes, granulocytes, or B cells in human blood. Cell association (%) refers to the proportion of each cell type with positive fluorescence, above background, stemming from fluorescence-labeled particles. Cell association (%) data are shown as the mean of three independent experiments (using the same batch of plasma from each donor), with at least 150,000 leukocytes analyzed for each experimental condition studied (see raw data in Figure S7). Change in cell association (%) refers to post-boost cell association (%) minus pre-vax cell association (%).

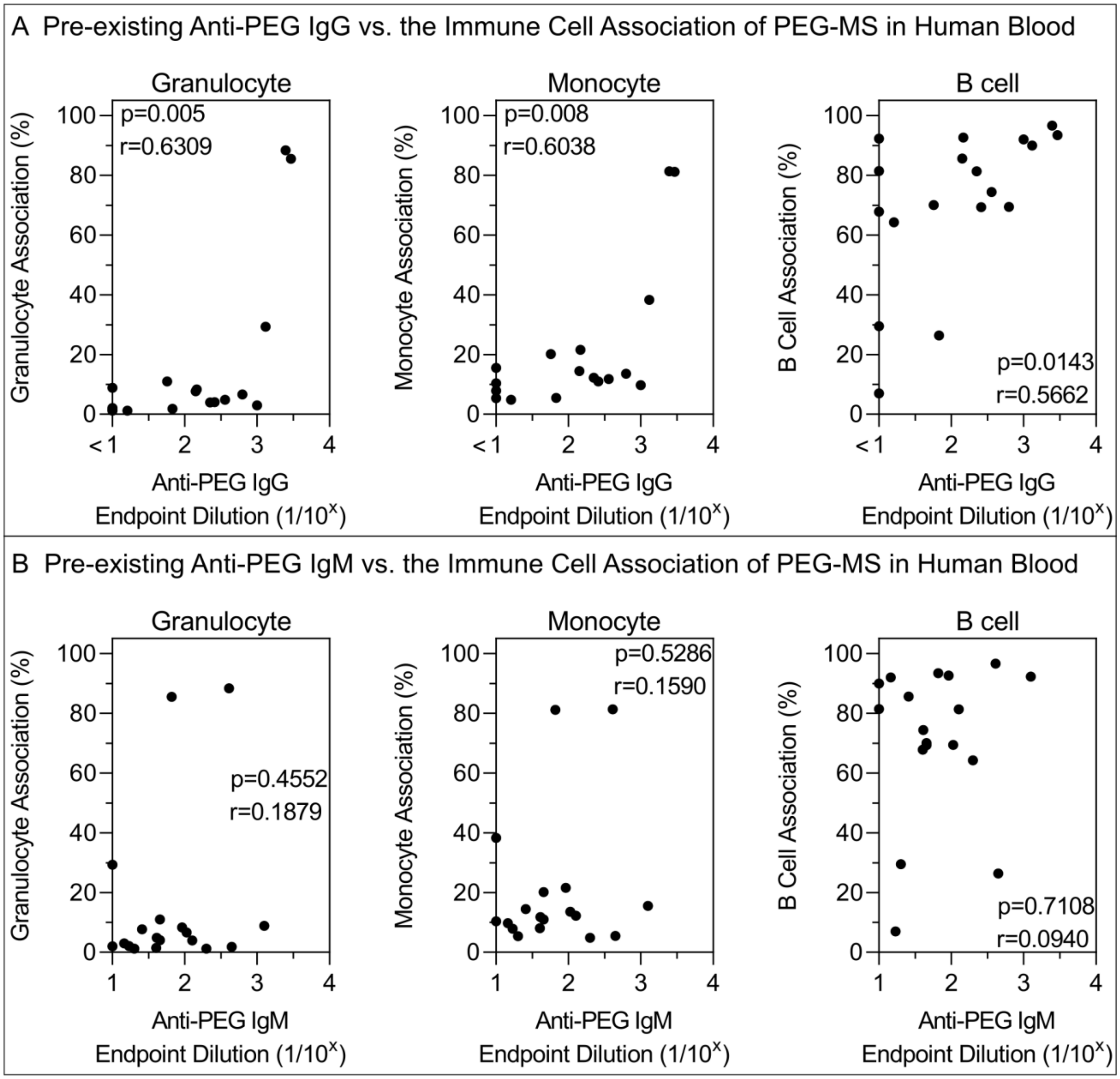


**Figure S11.** Impact of pre-existing anti-PEG antibody levels on the association of PEG-MS nanoparticles with immune cells in human blood. A,B) Spearman correlation between pre-existing (Pre-Vax) anti-PEG IgG or IgM titres (n=18, donor 1−18 from BNT162b2 vaccinee cohort) and PEG-MS nanoparticle association with monocytes, granulocytes, or B cells in human blood. Cell association (%) refers to the proportion of each cell type with positive fluorescence, above background, stemming from fluorescence-labeled particles. Cell association (%) data are shown as the mean of three independent experiments (using the same batch of plasma from each donor), with at least 150,000 leukocytes analyzed for each experimental condition studied (see raw data in Figure S8).

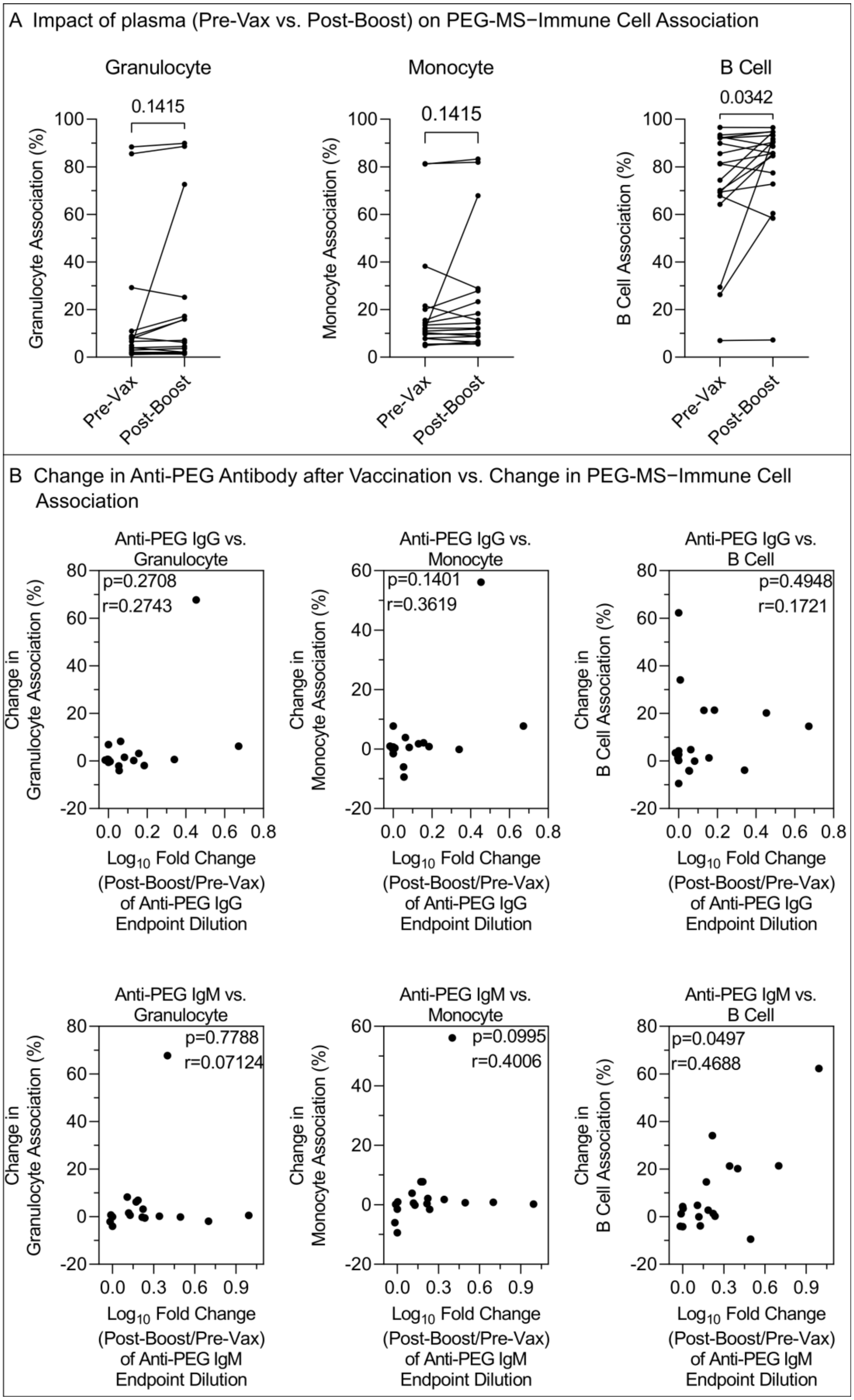
A Impact of plasma (Pre-Vax vs. Post-Boost) on PEG-MS−Immune Cell Association
Granulocyte
0.1415
Granulocyte Association (%)
Monocyte
0.1415
Monocyte Association (%)
B Cell
0.0342
B Cell Association (%)
Pre-Vax
Post-Boost
B Change in Anti-PEG Antibody after Vaccination vs. Change in PEG-MS−Immune Cell Association
Anti-PEG IgG vs. Granulocyte
p=0.2708
r=0.2743
Anti-PEG IgG vs. Monocyte
p=0.1401
r=0.3619
Anti-PEG IgG vs. B Cell
p=0.4948
r=0.1721
Change in Granulocyte Association (%)
Change in Monocyte Association (%)
Change in B Cell Association (%)
Log10 Fold Change (Post-Boost/Pre-Vax) of Anti-PEG IgG Endpoint Dilution
Anti-PEG IgM vs. Granulocyte
p=0.7788
r=0.07124
Anti-PEG IgM vs. Monocyte
p=0.0995
r=0.4006
Anti-PEG IgM vs. B Cell
p=0.0497
r=0.4688
Log10 Fold Change (Post-Boost/Pre-Vax) of Anti-PEG IgM Endpoint Dilution

**Figure S12.** Impact of change in anti-PEG antibody after BNT162b2 vaccination on the association of PEG-MS nanoparticles with immune cells in human blood. A) Comparing the impact of plasma collected before vaccination (Pre-Vax) and post-boost of the BNT162b2 vaccination (n=18, donor 1−18) on PEG-MS nanoparticle association with monocytes, granulocytes, or B cells in human blood. p-values were derived by Wilcoxon's matched-pairs signed rank test. B) Spearman correlation between the fold change ($log_{10}$) of anti-PEG IgG or IgM titres (Post-Boost/Pre-Vax) after 2 doses of BNT162b2 vaccination (n=18, donor 1−18) and change in PEG-MS nanoparticle association with monocytes, granulocytes, or B cells in human blood. Cell association (%) refers to the proportion of each cell type with positive fluorescence, above background, stemming from fluorescence-labeled particles. Cell association (%) data are shown as the mean of three independent experiments (using the same batch of plasma from each donor), with at least 150,000 leukocytes analyzed for each experimental condition studied (see raw data in Figure S8). Change in cell association (%) refers to post-boost cell association (%) minus pre-vax cell association (%).

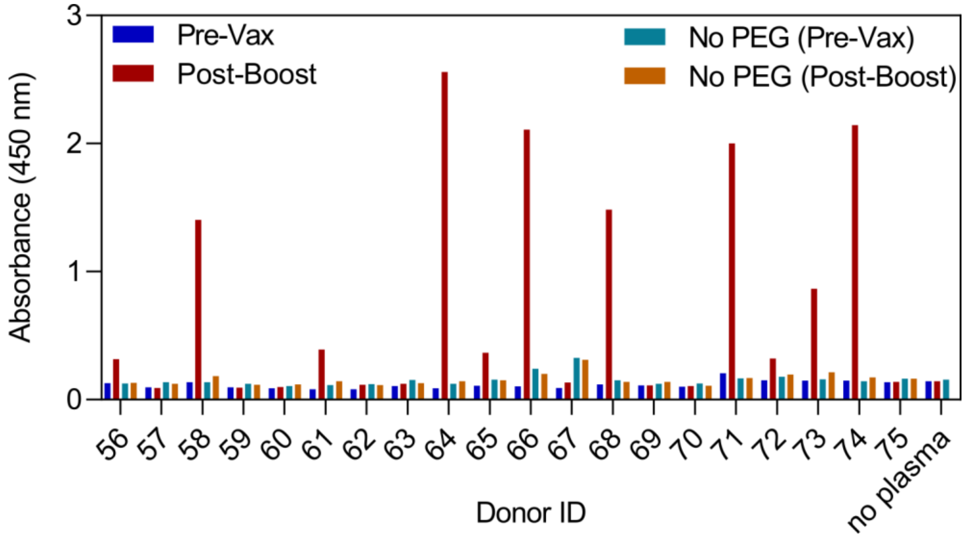


**Figure S13**. ELISA absorbance showing the PEG-specific antibody-mediated C1q binding of plasma collected before vaccination (Pre-Vax) and post-boost of the mRNA-1273 vaccination (n=20, donor 56−75). The ELISA were performed by coating 40kDa PEG on ELISA plates and the plasma were diluted at 1:5 in 1% BSA, followed by washing and incubating with Horseradish peroxidase (HRP)-conjugated C1q. No PEG-precoating wells were used as background. Data are shown as mean of two independent measurements.

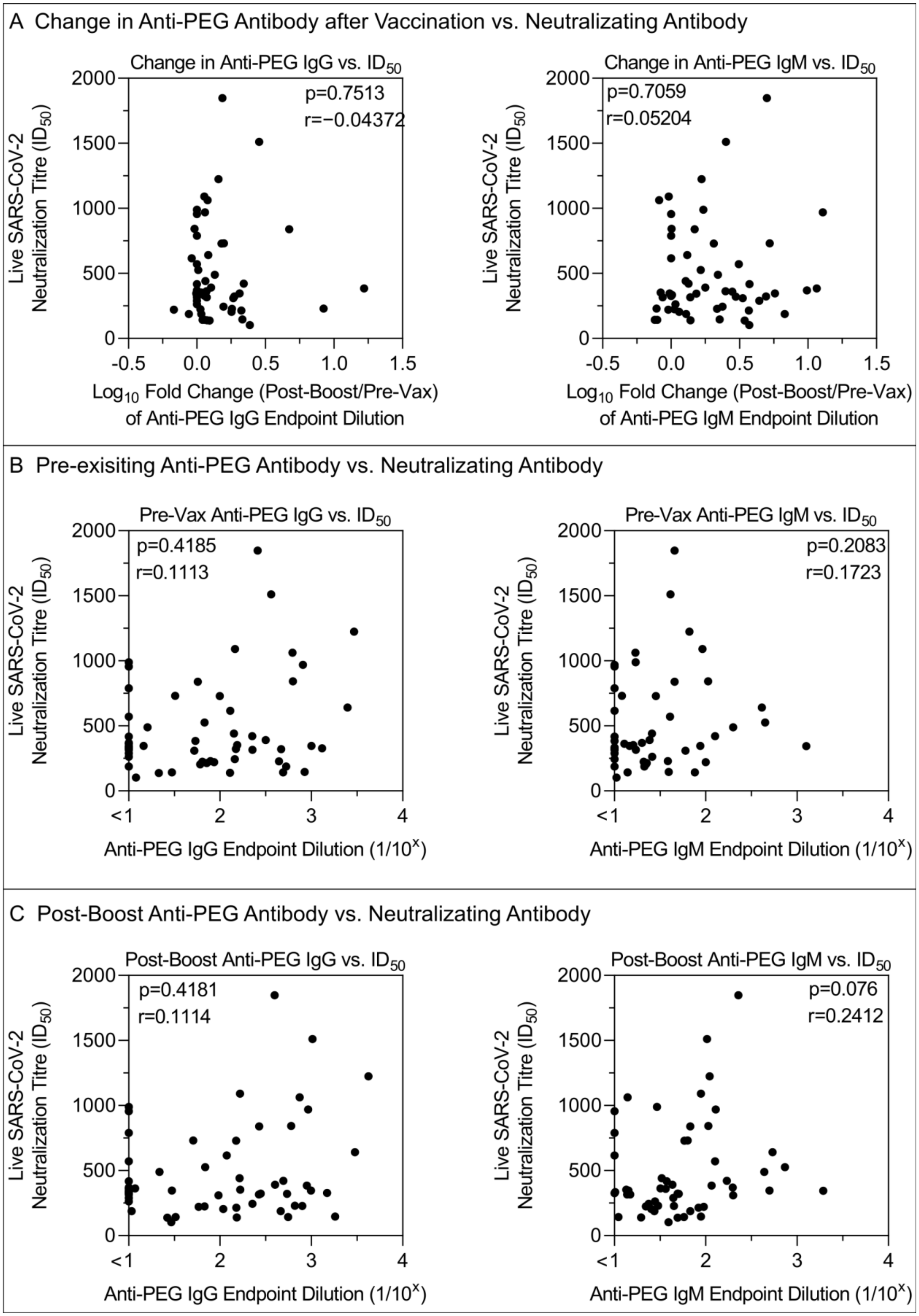
A Change in Anti-PEG Antibody after Vaccination vs. Neutralizating Antibody
Change in Anti-PEG IgG vs. ID50
p=0.7513
r=−0.04372
Live SARS-CoV-2 Neutralization Titre (ID50)
Log10 Fold Change (Post-Boost/Pre-Vax) of Anti-PEG IgG Endpoint Dilution
Change in Anti-PEG IgM vs. ID50
p=0.7059
r=0.05204
Log10 Fold Change (Post-Boost/Pre-Vax) of Anti-PEG IgM Endpoint Dilution
B Pre-exisiting Anti-PEG Antibody vs. Neutralizating Antibody
Pre-Vax Anti-PEG IgG vs. ID50
p=0.4185
r=0.1113
Anti-PEG IgG Endpoint Dilution (1/10^x)
Pre-Vax Anti-PEG IgM vs. ID50
p=0.2083
r=0.1723
Anti-PEG IgM Endpoint Dilution (1/10^x)
C Post-Boost Anti-PEG Antibody vs. Neutralizating Antibody
Post-Boost Anti-PEG IgG vs. ID50
p=0.4181
r=0.1114
Post-Boost Anti-PEG IgM vs. ID50
p=0.076
r=0.2412

**Figure S14**. The impact of PEG specific antibody on neutralizing antibody responses. A) Spearman correlation between live SARS-COV-2 neutralization titre (inhibitory dilution 50, $ID_{50}$) of post-boost plasma (donor 1-55 from BNT162b2 vaccinee cohort) and the fold change ($log_{10}$) of anti-PEG IgG or IgM titres (Post-Boost/Pre-Vax) after 2 dose of BNT162b2 vaccination (n=55, donor 1−55). B) Spearman correlation between live SARS-COV-2 neutralization titre (inhibitory dilution 50, $ID_{50}$) of post-boost plasma (donor 1-55 from BNT162b2 vaccinee cohort) and the pre-existing (Pre-Vax) anti-PEG IgG or IgM titres (n=55, donor 1−55 from BNT162b2 vaccinee cohort). C) Spearman correlation between live SARS-COV-2 neutralization titre (inhibitory dilution 50, $ID_{50}$) of post-boost plasma (donor 1-55 from BNT162b2 vaccinee cohort) and the post-boost anti-PEG IgG or IgM titres (n=55, donor 1−55 from BNT162b2 vaccinee cohort).

**Table S1.** BNT162b2 Vaccinee information

| Donor ID | Cohort ID | Sex | Age range | Days between two doses | Days post second dose sample collected[a] |
|---|---|---|---|---|---|
| 1 | 1 | F | 51-55 | 22 | 12 |
| 2 | 1 | M | 26-30 | 23 | 13 |
| 3 | 1 | F | 26-30 | 22 | 13 |
| 4 | 1 | F | 21-25 | 21 | 13 |
| 5 | 1 | M | 31-35 | 27 | 14 |
| 6 | 1 | F | 31-35 | 28 | 13 |
| 7 | 1 | F | 31-35 | 21 | 13 |
| 8 | 1 | F | 26-30 | 29 | 13 |
| 9 | 1 | F | 26-30 | 24 | 14 |
| 10 | 1 | F | 21-25 | 33 | 14 |
| 11 | 1 | M | 31-35 | 22 | 13 |
| 12 | 1 | F | 36-40 | 22 | 13 |
| 13 | 1 | M | 26-30 | 23 | 15 |
| 14 | 1 | F | 56-40 | 22 | 13 |
| 15 | 1 | F | 31-35 | 23 | 15 |
| 16 | 1 | M | 31-35 | 21 | 13 |
| 17 | 1 | F | 41-45 | 25 | 14 |
| 18 | 1 | M | 26-30 | 21 | 14 |
| 19 | 2 | F | 21-25 | 24 | 34 |
| 20 | 2 | F | 36-40 | 24 | 34 |
| 21 | 2 | F | 26-30 | 30 | 35 |
| 22 | 2 | M | 41-45 | 28 | 43 |
| 23 | 2 | M | 46-50 | 28 | 37 |
| 24 | 2 | M | 26-30 | 24 | 34 |
| 25 | 2 | F | 21-25 | 24 | 34 |
| 26 | 2 | F | 31-35 | 24 | 34 |
| 27 | 2 | M | 56-60 | 19 | 38 |
| 28 | 2 | F | 36-40 | 27 | 36 |
| 29 | 2 | F | 46-50 | 28 | 36 |
| 30 | 2 | F | 51-55 | 24 | 28 |
| 31 | 2 | F | 26-30 | 28 | 36 |
| 32 | 3 | F | 61-65 | 26 | 26/74 |
| 33 | 3 | M | 61-65 | 27 | 33/75 |
| 34 | 3 | M | 26-30 | 24 | 35/77 |
| 35 | 3 | F | 36-40 | 25 | 34/90 |
| 36 | 3 | M | 41-45 | 19 | 25/81 |
| 37 | 3 | F | 61-65 | 24 | 40/89 |
| 38 | 3 | F | 26-30 | 22 | 26/89 |
| 39 | 3 | M | 21-25 | 29 | 42/98 |
| 40 | 3 | M | 61-65 | 28 | 41/90 |

| | | | | | |
|---|---|---|---|---|---|
| 41 | 3 | M | 46-50 | 25 | 28/90 |
| 42 | 3 | F | 31-35 | 30 | 27 |
| 43 | 3 | M | 61-65 | 26 | 26 |
| 44 | 3 | F | 26-30 | 31 | 34 |
| 45 | 3 | F | 41-45 | 22 | 27 |
| 46 | 3 | F | 36-40 | 22 | 28 |
| 47 | 3 | F | 31-35 | 35 | 33 |
| 48 | 3 | F | 36-40 | 17 | 49 |
| 49 | 3 | M | 46-50 | 21 | 49 |
| 50 | 3 | F | 36-40 | 30 | 26 |
| 51 | 3 | F | 36-40 | 22 | 32 |
| 52 | 3 | M | 46-50 | 31 | 22 |
| 53 | 3 | F | 46-50 | 23 | 40 |
| 54 | 3 | M | 31-35 | 22 | 31 |
| 55 | 3 | F | 26-30 | 22 | 30 |
| Summary | | F: n=35 | Mean 38 (range 23−65) | Mean 25 (range 17−35) | 1st time point: Mean 27 (range 12−49) |
| | | M: n=20 | | | 2nd time point: Mean 85 (range 74−98) |

[a]Two post-boost time points were collected for donor 32−41, displayed as A/B.

**Table S2.** mRNA-1273 Vaccinee information

| Donor ID | Sex | Age range | Days between two doses | Days post second dose sample collected |
|---|---|---|---|---|
| 56 | F | 46-50 | 27 | 19 |
| 57 | M | 31-35 | 28 | 18 |
| 58 | F | 21-25 | 26 | 27 |
| 59 | F | 81-85 | 27 | 19 |
| 60 | M | 66-70 | 27 | 19 |
| 61 | M | 31-35 | 28 | 18 |
| 62 | F | 36-40 | 28 | 18 |
| 63 | M | 71-75 | 27 | 19 |
| 64 | F | 36-40 | 28 | 18 |
| 65 | M | 36-40 | 28 | 18 |
| 66 | F | 26-30 | 28 | 18 |
| 67 | F | 41-45 | 28 | 18 |
| 68 | F | 41-45 | 28 | 18 |
| 69 | M | 46-50 | 28 | 18 |
| 70 | M | 36-40 | 28 | 18 |
| 71 | F | 41-45 | 28 | 18 |
| 72 | F | 31-35 | 28 | 18 |
| 73 | M | 46-50 | 28 | 18 |
| 74 | M | 51-55 | 28 | 18 |
| 75 | F | 26-30 | 28 | 18 |
| Summary | F: n=11 | Mean 44 (range 24−84) | Mean 30 (range 26−28) | Mean 19 (range 18−27) |
| | M: n=9 | | | |

**Table S3.** Unvaccinated control blood donor information

| Donor ID | Sex | Age range | Health status[a] | Days in between two samples |
|---|---|---|---|---|
| 76 | F | 21-25 | COVID+ | 126 |
| 77 | F | 66-70 | COVID+ | 147 |
| 78 | M | 66-70 | COVID+ | 147 |
| 79 | M | 76-80 | COVID+ | 192 |
| 80 | F | 51-55 | COVID+ | 161 |
| 81 | M | 61-65 | COVID+ | 182 |
| 82 | M | 56-60 | COVID+ | 192 |
| 83 | F | 51-55 | COVID+ | 168 |
| 84 | F | 21-25 | COVID+ | 182 |
| 85 | F | 66-70 | COVID+ | 178 |
| 86 | F | 56-60 | COVID+ | 245 |
| 87 | M | 61-65 | COVID+ | 210 |
| 88 | M | 66-70 | COVID+ | 148 |
| 89 | M | 56-60 | COVID+ | 168 |
| 90 | F | 56-60 | COVID+ | 168 |
| 91 | M | 61-65 | COVID+ | 175 |
| 92 | M | 46-50 | COVID+ | 185 |
| 93 | F | 51-55 | COVID+ | 185 |
| 94 | M | 61-65 | COVID+ | 168 |
| 95 | M | 56-60 | COVID+ | 122 |
| 96 | F | 56-60 | COVID+ | 168 |
| 97 | F | 46-50 | COVID+ | 248 |
| 98 | M | 31-35 | COVID+ | 120 |
| 99 | F | 26-30 | COVID+ | 120 |
| 100 | F | 61-65 | COVID+ | 203 |
| 101 | M | 21-25 | COVID+ | 204 |
| 102 | M | 66-70 | COVID+ | 203 |
| 103 | M | 56-60 | COVID+ | 197 |
| 104 | F | 51-55 | COVID+ | 210 |
| 105 | F | 46-50 | COVID+ | 210 |
| 106 | M | 56-60 | COVID+ | 253 |
| 107 | F | 56-60 | COVID+ | 253 |
| 108 | M | 51-55 | COVID+ | 217 |
| 109 | F | 51-55 | COVID+ | 179 |
| 110 | M | 56-60 | COVID+ | 179 |
| 111 | F | 31-35 | COVID+ | 226 |
| 112 | M | 31-35 | COVID+ | 196 |
| 113 | M | 56-60 | COVID+ | 196 |
| 114 | F | 51-55 | COVID+ | 196 |
| 115 | F | 21-25 | COVID+ | 244 |

| | | | | |
|---|---|---|---|---|
| 116 | F | 51-55 | COVID- | 210 |
| 117 | M | 51-55 | COVID- | 210 |
| 118 | F | 56-60 | COVID- | 189 |
| 119 | F | 56-60 | COVID- | 196 |
| 120 | F | 56-60 | COVID- | 182 |
| 121 | F | 56-60 | COVID- | 182 |
| 122 | F | 21-25 | COVID- | 175 |
| 123 | F | 51-55 | COVID- | 175 |
| 124 | M | 51-55 | COVID- | 189 |
| 125 | F | 61-65 | COVID- | 193 |
| 126 | F | 61-65 | COVID- | 168 |
| 127 | F | 61-65 | COVID- | 168 |
| 128 | M | 61-65 | COVID- | 168 |
| 129 | M | 51-55 | COVID- | 175 |
| 130 | F | 51-55 | COVID- | 189 |
| Summary | F: n=31 | Mean 53 (range 21−76) | COVID+: 40 | Mean 186 (range 120−253) |
| | M: n=24 | | COVID-: 15 | |

[a]Donor 76−115 are SARS-CoV-2 infected convalescent subjects (COVID+) and Donor 116−130 are uninfected healthy donors (COVID-).

**Table S4.** Size and zeta potential of PEGylated nanoparticles

| Particle Type | Size (nm)[a] | Polydispersity Index[a] | Zeta Potential (mV)[b] |
|---|---|---|---|
| PEG-MS nanoparticles | 102 ± 30 | 0.015 | -32 ± 7 |
| Doxil | 72 ± 20 | 0.067 | -11 ± 9 |
| LNPs | 61 ± 18 | 0.079 | -10 ± 2 |

[a]The size and polydispersity index were determined by dynamic light scattering using a Zetasizer Nano-ZS (Malvern Instruments, UK) instrument.

[b]Zeta potential measurements were performed at pH 7.4 in phosphate buffer (5mM) using a Zetasizer Nano-ZS (Malvern Instruments, UK) instrument.

**Table S5**. Reported reactogenicity after boost of BNT162b2 vaccination from 44 donors

| **Donor ID:** | | **1** | **2** | **3** | **4** | **5** | **6** | **7** | **8** | **9** | **10** | **11** | **12** | **13** | **14** | **15** |
|---|---|---|---|---|---|---|---|---|---|---|---|---|---|---|---|---|
| **Injection Site** | Pain | Y | Y | Y | Y | Y | Y | Y | Y | Y | N | Y | Y | Y | Y | Y |
| | Redness | N | N | N | N | Y | N | Y | N | Y | N | N | Y | N | N | N |
| | Swelling | N | N | N | N | Y | N | N | N | Y | N | N | Y | N | N | Y |
| | Itching | N | N | N | N | N | N | N | N | N | N | N | N | N | N | Y |
| **Systemic** | Fatigue | N | Y | Y | N | N | Y | Y | Y | Y | Y | Y | Y | N | N | N |
| | Headache | N | N | Y | N | N | N | Y | N | N | Y | Y | N | N | N | Y |
| | Myalgia | N | Y | Y | N | N | N | N | Y | N | N | Y | N | N | N | N |
| | Chills | N | Y | Y | N | N | Y | Y | N | N | Y | N | N | N | N | Y |
| | Fever | N | N | Y | N | N | N | N | N | N | N | N | N | N | N | Y |
| | Joint pain | N | Y | Y | N | N | N | N | N | N | N | N | N | N | N | N |
| | Nausea | N | N | N | N | N | N | N | N | N | N | Y | N | N | N | N |
| | Vomiting | N | N | N | N | N | N | N | N | N | N | N | N | N | N | N |
| | Diarrhea | N | N | N | N | N | N | N | N | N | N | N | N | N | N | N |
| | Abdominal Pain | N | N | N | N | N | Y | N | N | N | N | N | N | N | N | N |
| | Rash (non-injection site) | N | N | N | N | N | N | N | N | N | N | N | N | N | N | N |
| Local Reactogenicity Score | | 1 | 1 | 1 | 1 | 3 | 1 | 2 | 1 | 3 | 0 | 1 | 3 | 1 | 1 | 3 |
| Systemic Reactogenicity Score | | 0 | 4 | 6 | 0 | 0 | 3 | 3 | 2 | 1 | 3 | 4 | 1 | 0 | 0 | 3 |
| Total Reactogenicity Score | | 1 | 5 | 7 | 1 | 3 | 4 | 5 | 3 | 4 | 3 | 5 | 4 | 1 | 1 | 6 |
| **Donor ID:** | | **16** | **17** | **18** | **19** | **20** | **21** | **22** | **23** | **24** | **25** | **26** | **27** | **28** | **29** | **30** |
| **Injection Site** | Pain | N | N | Y | N | Y | Y | Y | Y | Y | Y | Y | Y | Y | Y | N |
| | Redness | N | N | N | Y | N | N | Y | Y | Y | Y | Y | Y | Y | Y | N |
| | Swelling | Y | N | Y | Y | N | N | N | N | N | N | N | N | N | N | N |
| | Itching | N | N | N | N | N | N | N | N | N | N | N | N | N | N | N |
| **Systemic** | Fatigue | Y | Y | Y | N | N | N | Y | N | Y | Y | Y | Y | N | N | N |
| | Headache | Y | N | Y | N | N | Y | N | N | N | N | Y | N | N | Y | N |
| | Myalgia | Y | N | Y | N | N | Y | N | N | Y | Y | N | Y | N | N | N |
| | Chills | N | N | Y | N | N | N | N | N | N | N | N | N | N | N | N |
| | Fever | Y | Y | Y | N | Y | N | N | Y | N | N | Y | N | N | N | N |

| | | | | | | | | | | | | | | | | |
|---|---|---|---|---|---|---|---|---|---|---|---|---|---|---|---|---|
| | Joint pain | Y | N | N | N | N | N | N | N | N | Y | N | Y | N | N | N |
| | Nausea | N | N | N | N | Y | N | N | N | N | Y | N | N | N | N | N |
| | Vomiting | N | N | N | N | N | N | N | N | N | N | N | N | N | N | N |
| | Diarrhea | N | N | N | N | N | N | N | N | N | N | N | N | N | N | N |
| | Abdominal Pain | N | N | N | N | N | N | N | N | N | N | N | N | N | N | N |
| | Rash (non-injection site) | N | N | N | N | N | N | N | N | N | N | N | N | N | N | N |
| Local Reactogenicity Score | | 1 | 0 | 2 | 2 | 1 | 1 | 2 | 2 | 2 | 2 | 2 | 2 | 2 | 2 | 0 |
| Systemic Reactogenicity Score | | 5 | 2 | 5 | 0 | 2 | 2 | 1 | 1 | 2 | 4 | 3 | 3 | 0 | 1 | 0 |
| Total Reactogenicity Score | | 6 | 2 | 7 | 2 | 3 | 3 | 3 | 3 | 4 | 6 | 5 | 5 | 2 | 3 | 0 |
| **Donor ID:** | | **31** | **32** | **33** | **40** | **41** | **42** | **43** | **45** | **47** | **50** | **51** | **53** | **54** | **55** | |
| **Injection Site** | Pain | N | Y | Y | N | N | Y | Y | Y | N | N | Y | N | Y | N | |
| | Redness | N | N | N | N | N | N | Y | Y | N | N | N | N | N | N | |
| | Swelling | N | N | N | N | N | N | N | Y | N | N | N | N | N | N | |
| | Itching | N | N | N | N | N | N | N | Y | N | N | N | N | N | N | |
| **Systemic** | Fatigue | N | N | N | N | N | N | N | Y | N | N | N | Y | N | Y | |
| | Headache | N | N | N | Y | N | Y | N | Y | N | N | N | N | N | Y | |
| | Myalgia | N | N | N | N | N | N | N | N | N | N | N | Y | N | Y | |
| | Chills | N | N | N | N | N | N | N | N | N | N | N | Y | N | Y | |
| | Fever | N | N | N | N | N | N | N | N | N | N | N | N | N | Y | |
| | Joint pain | N | N | N | N | N | N | N | N | N | N | N | Y | N | N | |
| | Nausea | N | N | N | N | N | N | N | N | N | N | N | N | N | Y | |
| | Vomiting | N | N | N | N | N | N | N | N | N | N | N | N | N | N | |
| | Diarrhea | N | N | N | N | N | N | Y | N | N | N | N | N | N | N | |
| | Abdominal Pain | N | N | N | N | N | N | N | N | N | N | N | N | N | N | |
| | Rash (non-injection site) | N | N | N | N | N | N | N | N | N | N | N | N | N | N | **Mean** |
| Local Reactogenicity Score | | 0 | 1 | 1 | 0 | 0 | 1 | 2 | 4 | 0 | 0 | 1 | 0 | 1 | 0 | 1.3 |
| Systemic Reactogenicity Score | | 0 | 0 | 0 | 1 | 0 | 1 | 1 | 2 | 0 | 0 | 0 | 4 | 0 | 6 | 1.7 |
| Total Reactogenicity Score | | 0 | 1 | 1 | 1 | 0 | 2 | 3 | 6 | 0 | 0 | 1 | 4 | 1 | 6 | 3.0 |

**Table S6**. Reported reactogenicity after boost of mRNA-1273 vaccination from 19 donors

| **Donor ID:** | | **56** | **57** | **58** | **59** | **60** | **61** | **62** | **63** | **64** | **65** |
|---|---|---|---|---|---|---|---|---|---|---|---|
| **Injection Site** | Pain | Y | Y | Y | Y | Y | Y | Y | N | Y | N |
| | Redness | Y | N | N | N | N | N | N | N | N | Y |
| | Swelling | Y | N | Y | N | N | Y | N | N | N | N |
| | Itching | N | N | N | N | N | Y | N | N | N | N |
| **Systemic** | Fatigue | Y | Y | N | N | Y | N | Y | N | Y | N |
| | Headache | Y | N | Y | N | Y | N | Y | N | N | N |
| | Myalgia | Y | Y | N | N | Y | Y | N | N | Y | Y |
| | Chills | Y | N | Y | N | N | N | Y | N | Y | Y |
| | Fever | Y | N | Y | N | N | N | Y | N | Y | Y |
| | Joint pain | Y | Y | Y | N | Y | N | N | N | N | Y |
| | Nausea | Y | N | N | N | N | N | Y | N | N | N |
| | Vomiting | N | N | N | N | N | N | N | N | N | N |
| | Diarrhea | N | N | Y | N | N | N | N | N | N | N |
| | Abdominal Pain | N | N | N | N | N | N | Y | N | N | N |
| | Rash (non-injection site) | N | N | N | N | N | N | N | N | N | N |
| Local Reactogenicity Score | | 3 | 1 | 2 | 1 | 1 | 3 | 1 | 0 | 1 | 1 |
| Systemic Reactogenicity Score | | 7 | 3 | 5 | 0 | 4 | 1 | 6 | 0 | 4 | 4 |
| Total Reactogenicity Score | | 10 | 4 | 7 | 1 | 5 | 4 | 7 | 0 | 5 | 5 |
| **Donor ID:** | | **66** | **67** | **68** | **69** | **70** | **71** | **73** | **74** | **75** | |
| **Injection Site** | Pain | N | Y | Y | Y | Y | Y | Y | Y | Y | |
| | Redness | N | N | Y | N | N | N | N | N | Y | |
| | Swelling | N | N | Y | Y | N | N | N | N | N | |
| | Itching | N | N | N | N | N | N | N | N | N | |
| **Systemic** | Fatigue | N | Y | Y | Y | Y | Y | Y | Y | Y | |
| | Headache | N | Y | Y | N | Y | Y | Y | Y | Y | |
| | Myalgia | N | Y | Y | Y | Y | Y | Y | Y | Y | |
| | Chills | N | N | Y | Y | Y | Y | Y | Y | Y | |
| | Fever | N | N | Y | N | Y | N | Y | Y | N | |
| | Joint pain | N | Y | N | Y | Y | Y | Y | N | Y | |
| | Nausea | N | N | N | N | N | Y | N | Y | Y | |
| | Vomiting | N | N | N | N | N | Y | N | N | N | |

| | | | | | | | | | | | |
|---|---|---|---|---|---|---|---|---|---|---|---|
| | Diarrhea | N | N | N | N | N | Y | N | N | N | |
| | Abdominal Pain | N | N | N | N | N | Y | N | N | N | |
| | Rash (non-injection site) | N | N | N | N | N | N | N | N | N | **Mean** |
| Local Reactogenicity Score | | 0 | 1 | 3 | 2 | 1 | 1 | 1 | 1 | 2 | 1.4 |
| Systemic Reactogenicity Score | | 0 | 4 | 5 | 4 | 6 | 9 | 6 | 6 | 6 | 4.2 |
| Total Reactogenicity Score | | 0 | 5 | 8 | 6 | 7 | 10 | 7 | 7 | 8 | 5.6 |
